# Pathogenic variants in *BORCS5* Cause a Spectrum of Neurodevelopmental and Neurodegenerative Disorders with Lysosomal Dysfunction

**DOI:** 10.1101/2025.04.30.25326597

**Authors:** Niccolò E. Mencacci, Georgia Minakaki, Reza Maroofian, Raffaella De Pace, Adeline Paimboeuf, Patrick Shannon, David Chitayat, Francesca Magrinelli, Wesley J. Peng, Diptaman Chatterjee, Sara H. Eldessouky, Julia Baptista, Tamas Marton, Julie Vogt, Juan Dario Ortigoza-Escobar, Loreto Martorell, Marta Gómez-Chiari, Ingrid M Wentzensen, Erik-Jan Kamsteeg, Maha S. Zaki, Annarita Scardamaglia, Giovanni Zifarelli, Zuhair Nasser Al-Hassnan, Elka Miller, Shiri Shinar, Lova S. Matsa, Sri Hari Chandan Appikonda, Michael Schwake, Mariasavina Severino, Henry Houlden, Shunmoogum A. Patten, Juan S. Bonifacino, Kailash P. Bhatia, Dimitri Krainc

## Abstract

*BORCS5* encodes a subunit of the BLOC-one-related complex (BORC), which is known to mediate the kinesin-dependent anterograde movement of lysosomes. Using whole-exome sequencing, we identified 12 cases from seven families carrying bi-allelic *BORCS5* variants, including four loss-of-function and two missense variants. Carriers of homozygous loss-of-function variants presented with prenatally lethal arthrogryposis multiplex congenita, brain malformations, and neuropathological evidence of diffuse neuroaxonal dystrophy. Individuals with missense variants presented differently, with microcephaly, developmental epileptic encephalopathy, intellectual disability, optic atrophy, spasticity, and progressive movement disorders. In this group, brain MRI showed diffuse hypomyelination and progressive global cerebral atrophy, consistent with neurodegeneration. *Borcs5* knockout in zebrafish exhibited microcephaly, motor deficits, and seizures, mirroring the patients’ clinical presentation. At the cellular level, BORCS5 loss-of-function but not missense variants, resulted in lower protein expression and impaired BORC assembly, paralleled by perinuclear lysosomal clustering. However, both loss-of-function and missense BORCS5 variants were associated with reduced total lysosomal proteolysis, reduced activity of the lysosomal hydrolases glucocerebrosidase and cathepsin B, and presence of multilamellar bodies, indicating lysosomal dysfunction. Our study reveals a novel role for BORCS5 in the regulation of lysosomal function, in addition to its known role in the anterograde movement of lysosomes, possibly underlying the diverse clinical manifestations in individuals with BORCS5-related disorders.

## Introduction

Genetic and functional studies have implicated rare and common variants in multiple lysosomal genes in the pathogenesis of various neurological disorders, such as Parkinson’s disease (PD)^1,2^ and dystonia^3,4^. Identifying and characterizing rare genetic causes of lysosomal dysfunction facilitates the discovery of molecular modifiers of lysosomal activity and development of targeted therapies in neurodegeneration. Pathogenic variants can affect various aspects of lysosomal biology, including the cellular processes of lysosomal maturation, fusion with late endosomes or autophagosomes, post-translational modifications of lysosomal components, or trafficking of lysosomal hydrolases from the endoplasmic reticulum/trans-Golgi network.^5–7^

*BORCS5* (MIM *616598; previously known as *LOH12CR1*) encodes the 196-amino-acid protein <u>BLOC</u>-<u>O</u>ne-<u>R</u>elated <u>C</u>omplex subunit 5 (BORCS5; also known as Myrlysin), a subunit of the <u>BLOC</u>-<u>O</u>ne-<u>R</u>elated <u>C</u>omplex (BORC). BORC is a ubiquitously expressed protein complex composed of eight subunits named BORCS1–8,^8^ which is known for mediating the interaction between lysosomes and kinesins via ARL8b and its effector SKIP, allowing for microtubule-mediated anterograde trafficking of lysosomes.^9,10^ In neurons, BORC controls anterograde movements of lysosomes particularly in axons,^11–13^ but the consequences of disrupted axonal transport of lysosomes in neurological disorders are not fully understood.

It was recently shown that BORC also mediates axonal transport of RNA granules hitchhiking on lysosome-related structures.^14^ Disruption of BORC resulted in depletion of axonal mRNAs encoding proteins essential for mitochondrial function.^14^ Additional functions that have been attributed to BORC include autophagosome-lysosome fusion via the HOPS complex^15^ and maturation of late endosome to lysosome.^16^ Loss of either Borcs5 or Borcs7 in mouse models results in clustering of lysosomes in the soma and their depletion from axons, leading to axonal pathology, motor dysfunction and perinatal respiratory failure.^11,13^ Moreover, bi-allelic pathogenic variants in *BORCS8* were recently identified in three unrelated families with an infantile-onset neurodegenerative disorder (MIM# 620987).^17^

We report the identification of bi-allelic pathogenic variants in *BORCS5,* including both loss-of-function (LoF) and missense variants, in 12 subjects from seven unrelated families. The clinical presentation of these cases ranged from perinatally lethal arthrogryposis multiplex congenita (AMC) associated with brain malformations and neuropathological evidence of neuroaxonal dystrophy (NAD), to a severe neurodegenerative phenotype with developmental epileptic encephalopathy and subsequent progressive movement disorders. Cellular studies showed that only *BORCS5* LoF variants resulted in perinuclear lysosomal clustering, whereas both LoF and missense *BORCS5* variants were associated with multiple features of lysosomal dysfunction. Our data suggest a dual role for BORCS5 in both lysosomal anterograde trafficking and lysosomal function. The impact of *BORCS5* variants on different aspects of lysosomal biology may underlie the different clinical presentations observed in individuals with *BORCS5*-related neurological disorders.

## Results

### Identification of biallelic BORCS5 variants in affected subjects from seven unrelated families

Seven unrelated families were recruited into the study (Fig. 1A). In total, we identified six pathogenic variants in *BORCS5* (NM_058169.4), including two missense variants (c.284G>A, p.R95Q; c.296A>C, p.H99P) and four LoF alleles (c.203-1G>T, p.?; c.316delG, p.A106Pfs*20; c.382_383delAG, p.L128Vfs*86; c.417C>G, p.Y139*) for a total of 12 affected subjects (Fig. 1A, 1B).

**Figure 1.**
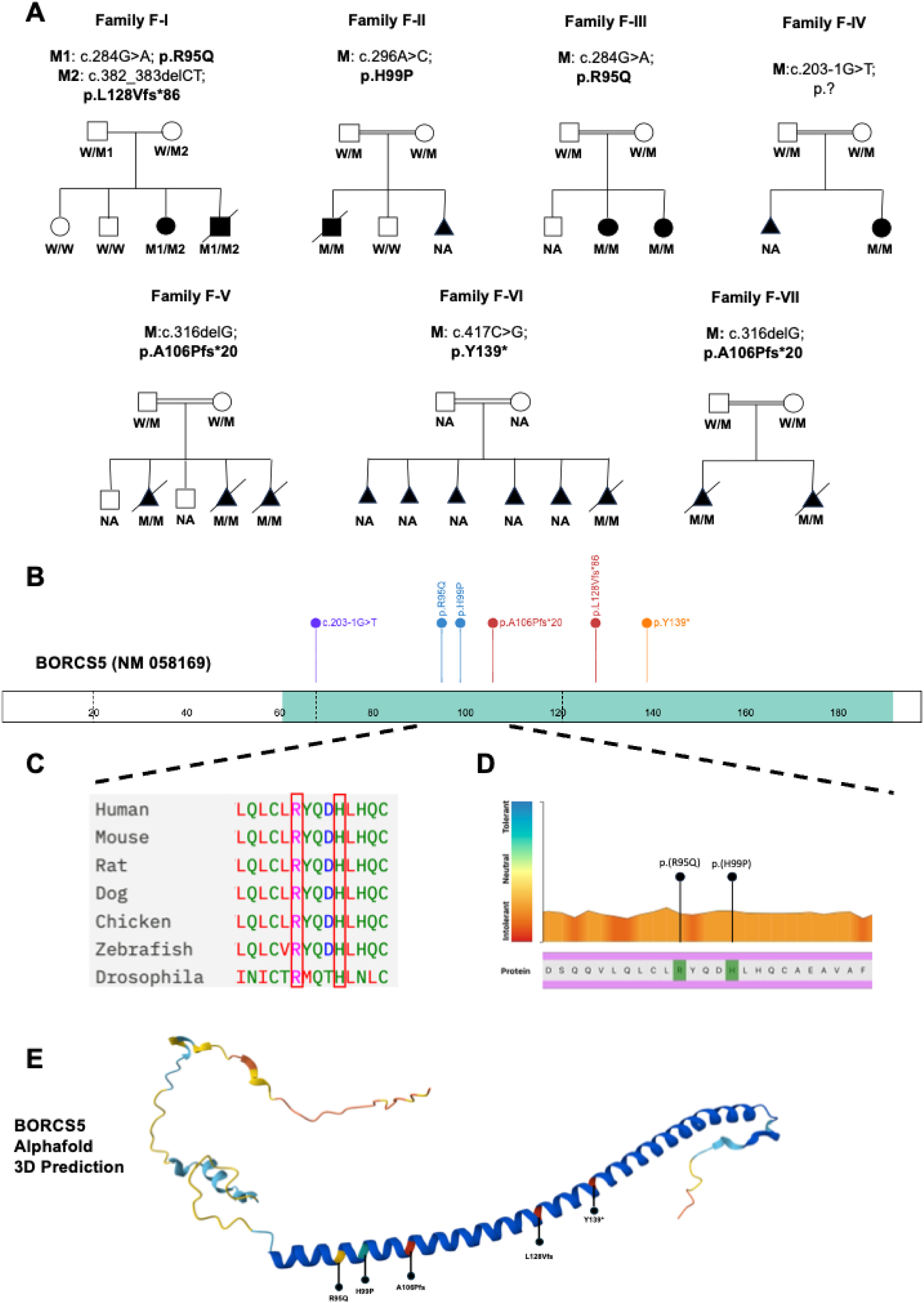
Family pedigrees and bi-allelic *BORCS5* variants. A. Family pedigrees and genotypes of cases with bi-allelic *BORCS5* variants. Triangles indicate spontaneous miscarriages, while crossed triangles indicate elective pregnancy terminations. B. Schematic of BORCS5 protein indicating the position of the identified pathogenic *BORCS5* variants. C. Conservation across species of the amino acid residues involved by the identified pathogenic missense variants R95Q and H99P. D. Graphic representation of intolerance to *BORCS5* variants. Using the Metadome software (https://stuart.radboudumc.nl/metadome/), we mapped the identified missense variants, which both affect amino acid residues that show significant intolerance to their variation. E: Structure of BORCS5 predicted by AlphaFold and localization of coding variants identified in this study.

In the index pedigree (family F-I), quad familial research whole-exome sequencing (WES) of DNA from the two affected siblings (F-I:1 and F-I:2) and their healthy parents identified two rare variants in *trans* in *BORCS5*, the maternally inherited frameshift variant L128Vfs*86 and the paternally inherited missense variant R95Q. Sanger sequencing confirmed the variants and showed that the two older unaffected siblings were not carriers.

10 additional cases from six families with *BORCS5* bi-allelic variants were subsequently identified. Based on segregation analysis, *BORCS5* variants were independently prioritized as the top candidate genetic etiology in each family.

Only affected family members from F-I had compound heterozygous *BORCS5* variants, whereas all other affected subjects carried homozygous variants. The two affected cases from family F-III carried the homozygous missense variant R95Q, which was also found in family F-I. The same homozygous LoF variant A106Pfs*20 was identified in 5 cases from two unrelated Pakistani families (F-V and F-VII). The homozygous variants H99P (F-II:1), c.203-1G>T (F-IV:1), and Y139* (F-VI:1), were identified in single individual cases.

All identified *BORCS5* variants (see Supplementary Table 1) are absent or exceedingly rare in gnomAD (minor allele frequency < 0.000001). Additionally, there are no entries for homozygous LoF variants in gnomAD.

The two missense variants R95Q and H99P have CADD scores > 20, which indicates these variants are in the top 1% most deleterious changes for the human genome and are predicted to be damaging by all available *in silico* tools. Furthermore, both variants affect amino acid residues that are completely conserved across species and are localized in a portion of the protein that is highly intolerant to genetic variation (Fig. 1C, 1D).

The AlphaFold prediction of the BORCS5 protein structure indicates that amino acid residues R95 and H99 are part of a long α-helix which is likely to be essential for its interaction with other BORC subunits (Fig. 1E). The LoF variants A106Pfs* and Y139* are predicted to cause an early protein truncation, removing a significant proportion of the BORCS5 α-helical domain. The L128Vfs allele is predicted to lead to a profound alteration in the amino acid composition of the C-terminus of BORCS5, extending the protein by 16 amino acids and likely resulting in a severe disruption of the protein α-helix. The splice-site acceptor variant c.203-1G>T is predicted to result in a complete loss of the acceptor site according to Splice AI.

### BORCS5 bi-allelic variants cause a phenotypic spectrum ranging from severe developmental epileptic encephalopathy with neurodegeneration and progressive movement disorders to perinatal lethality with brain malformations and neuroaxonal dystrophy

We describe a total of 12 affected subjects with bi-allelic *BORCS5* variants.

Six individuals from families F-I, F-II, F-III, and F-IV exhibited a core phenotype of developmental encephalopathy with severe intellectual disability, delayed milestones, seizures, and progressive movement disorders with spasticity. Most had the *BORCS5* missense variants R95Q or H99P, either in compound heterozygosity with a LoF variant (F-I) or in homozygosity (F-II and F-III). The case from F-IV, which was previously reported with minimal clinical details,^18^ carried the splice-site acceptor variant c.203-1G>T, p.(?).

None of these cases attained speech or walking abilities, requiring complete dependence. Developmental regression, including loss of babbling, visual tracking, head control, and ability to stand, was observed in the three cases from F-I and F-II. Seizures occurred within the first year, managed effectively with standard antiepileptic treatments in families F-I, F-II, and F-III. Older individuals from F-I became seizure-free in their teens. However, case F-IV:1 had a more severe epileptic phenotype, featuring infantile spasms and an EEG showing hypsarrhythmia. Seizures evolved into myoclonic seizures, poorly controlled with Vigabatrin, Levetiracetam, Topiramate, and Clonazepam but partially responsive to ketogenic diet.

All cases exhibited progressive spasticity, diffuse hyperreflexia, bilateral extensor plantar response, and severe limb contractures. Movement disorders (dystonia and parkinsonism) were prominent in families F-I and F-II (see Supplementary Video). Generalized dystonia led to scoliosis, painful spasms, and oculogyric crises. Parkinsonian features included severe limb bradykinesia and hypomimia. Dystonic spasms improved dramatically with carbidopa/levodopa, suggesting nigrostriatal dopaminergic denervation. Other medications, including trihexyphenidyl, baclofen, and diazepam, helped to manage spasticity and dystonia.

Optic neuropathy was present in all subjects. Nerve conduction studies revealed sensorimotor demyelinating neuropathy in F:II-1 but were normal in F:I-1. Dysmorphic features included microcephaly, dolichocephaly, and low-set ears. Swallowing difficulties necessitated percutaneous endoscopic gastrostomy in cases from families F-I and F-II.

Neuroimaging in five cases from F-I, F-II, and F-III showed cerebral atrophy, white matter loss, hypomyelination, small T2-hypointense thalami, thin brainstem, and optic nerve atrophy (Fig. 2). F-I:1 and F-II:1 also had mild cerebellar atrophy. Changes in F-II:1 progressed rapidly within the first. 2 years of life, indicating an aggressive neurodegenerative process (Fig. 2C-D). Conversely, case F-IV:1 revealed a different pattern characterized by corpus callosum agenesis and polymicrogyria.

**Figure 2.**
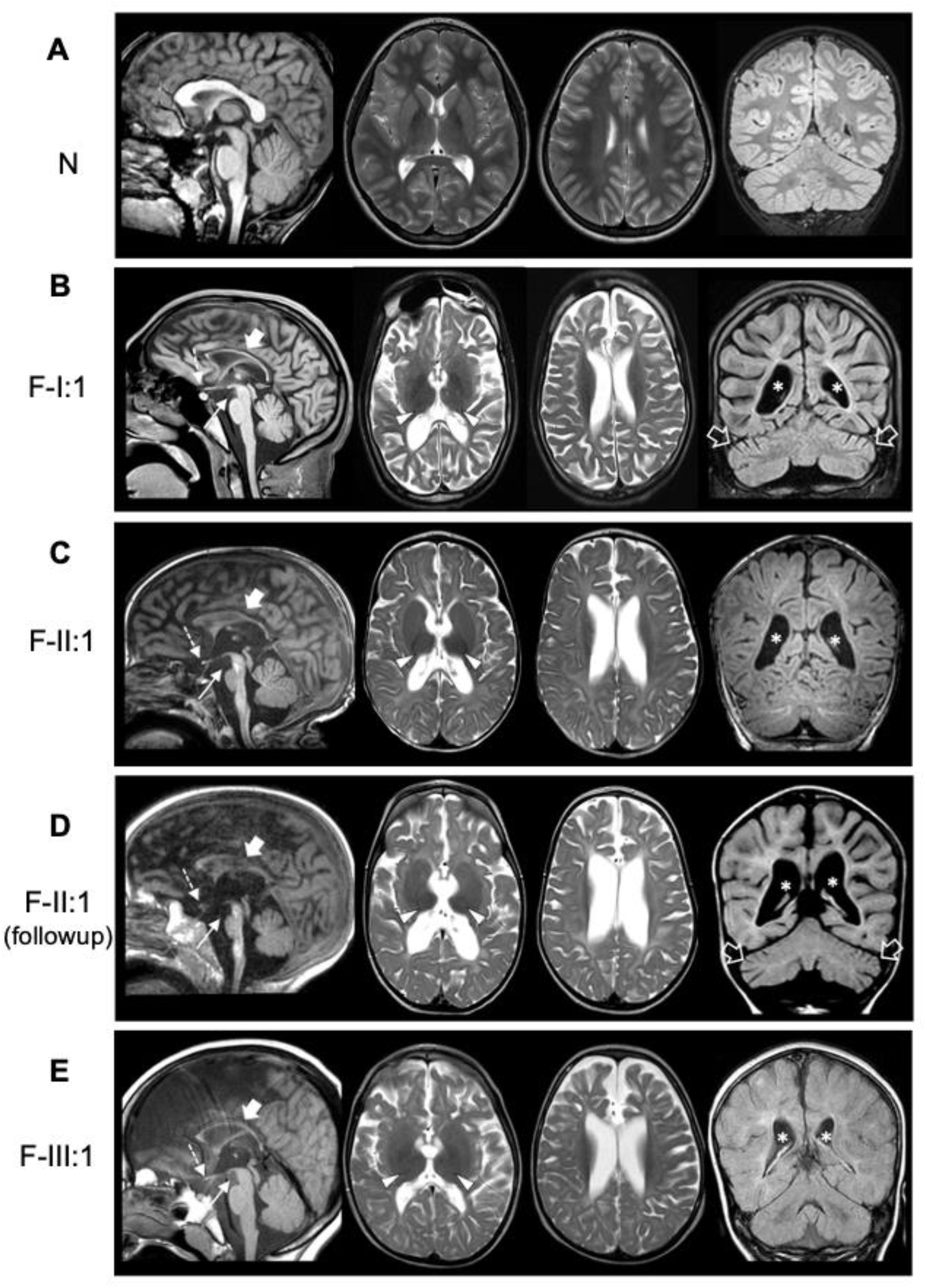
Neuroimaging findings in cases with *BORCS5* missense variants. Brain MRI studies of a control subject (A) for comparison individuals F-I:1 (B), F-II:1 in the first year of life (C) and 1 year later (D), and F-III:1 (E); sagittal T1-weighted images (first column), axial T2-weighted images (second and third column), and coronal FLAIR or T1-weighted images (last column). There is moderate to severe cerebral atrophy and loss of white matter volume with consequent ventricular dilatation in all subjects (asterisks). The myelination is markedly reduced/incomplete in all cases. The corpus callosum is very thin in all individuals (thick arrows) with associated hypoplastic anterior commissure. There is atrophy of the optic nerves (not shown) and chiasm (dashed arrows) in all patients. The thalami are small and hypointense on T2-weighted images (arrowheads). The midbrain and pons are small in all individuals (thin arrows), especially FII:1. Mild atrophy is noted in F-I:1 and F-II:1 (empty arrows). Note the clear progression of cerebral and cerebellar atrophy with arrested myelination in FII:1.

Six cases from families F-V, F-VI, and F-VII with biallelic *BORCS5* LoF early truncating variants (A106Pfs or Y139*) displayed a distinct phenotype with severe prenatal neurological manifestations, initially manifesting with reduced fetal movements. Intrauterine imaging studies revealed in all cases AMC, hydrocephalus with aqueduct stenosis, agenesis of the corpus callosum, markedly thin brainstem, cerebellar hypoplasia, and diffuse muscle atrophy (Supplementary Fig. 1A). Growth parameters and other organ systems were normal. Pregnancies of these cases were all terminated between 21 and 25 weeks of gestation.

Histopathological examination of the brain from two fetuses from families F-V and F-VII (Fig. 3, Supplementary Fig. 1) revealed microcephaly, hypoplastic thalamus, basal ganglia and cerebellum, alongside ventriculomegaly, aqueductal atresia, and absent corpus callosum (Fig. 3A, 3B). Examination of the brainstem showed hypoplasia of major neuronal groups, including tegmental nuclei, inferior olivary nuclei, basis pontis, and dentate nucleus (Fig. 3C, 3D, 3E) Furthermore, histological analysis revealed in both cases widespread neuroaxonal degeneration with formation of axonal spheroids, particularly in the brainstem, cerebral hemispheres, and peripheral roots and nerves (Fig. 3F, 3I, Supplementary Fig. 1E, 1G). Immunohistochemistry revealed strong β-amyloid precursor protein, neurofilament, and α-synuclein staining of the axonal spheroids, consistent with a pathological diagnosis of NAD (Fig. 3G, 3H, 3J; Supplementary Fig. 1B, 1F). However, axonal spheroids did not stain positive for phosphorylated α-synuclein (Supplementary Fig. 1C), tau, or TDP-43.

**Figure 3.**
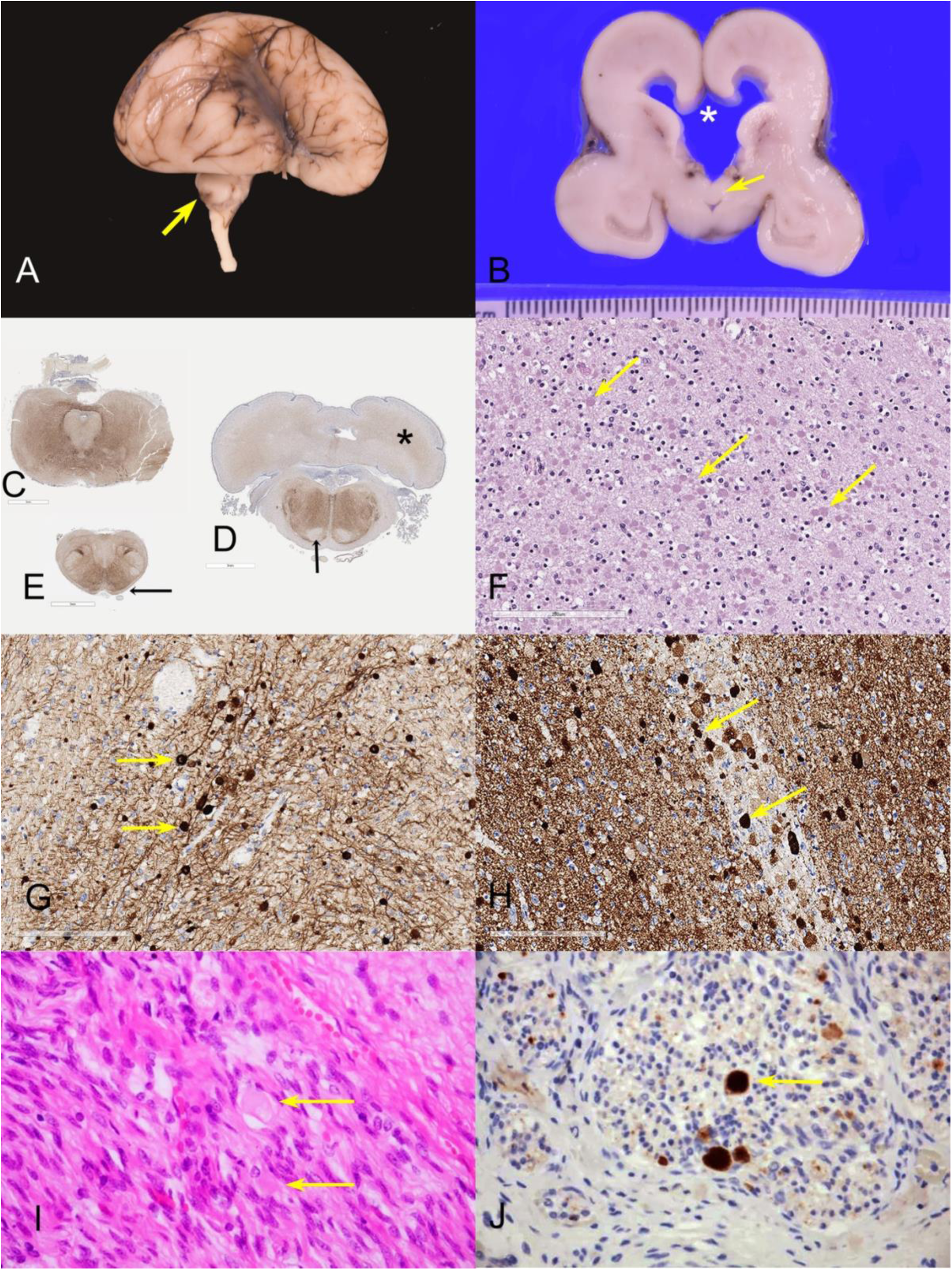
Pathological characterization of cases with bi-allelic loss-of-function *BORCS5* variants. A: Brain of case F-VII:1 demonstrating hypoplastic temporal lobes, a delayed, smooth Sylvian fissure and a markedly hypoplastic cerebellum (arrow), B. Coronal section, demonstrating ventriculomegaly. The corpus callosum is reduced to a thin membrane and has ruptured (asterisk). The septum is ruptured, and the fornices (arrow) are descended and lie on the roof of the third ventricle. C-D-E: Immunohistochemistry for neurofilament light chain in whole mounts of posterior fossa structures showing: midbrain with minute aqueduct and hypoplastic cerebral peduncles (C); the caudal pons with very small and smooth inferior olivary nuclei, hypoplastic middle cerebellar peduncles, absent corticofugal tracts (arrow) and poorly defined dentate nuclei (asterisk) (D); absent pyramids (arrows) and inferior cerebellar peduncles in the medulla (E). F-G-H: Histological analysis of the same case with hematoxylin and eosin and immunohistochemistry demonstrating innumerable pale eosinophilic axonal spheroids (arrows) in the internal capsule (F); strong positive staining of the axonal spheroids for neurofilament light chain (G) and alpha-synuclein (H). I-J: Peripheral nerves of Case F-V:2 demonstrated numerous axonal spheroids (arrows) (I), which stained strongly positive for β-amyloid precursor protein (J).

### Zebrafish borcs5 knockout leads to neurodevelopmental defects

To investigate the effect of loss of BORCS5 *in vivo*, we knocked out *borcs5* in zebrafish (borcs5-ko) using CRISPR-Cas9 editing to induce targeted biallelic *borcs5* mutations directly in the injected embryos (F0 generation).^17,19^ The zebrafish genome encodes a single *borcs5* ortholog, sharing 84% amino acid identity with human *BORCS5*.

At 3 days post-fertilization (3 dpf), borcs5-ko larvae exhibited reduced body size with a slight curvature of the body axis (Fig. 4A, 4B). Importantly, borcs5-ko larvae had reduced eye and head size relative to WT (Fig. 4C, 4D), and these gross morphological defects persisted at 5 dpf (Supplementary Fig. 2C–F). Injection of human *BORCS5^WT^* mRNA in borcs5-ko zebrafish rescued morphological phenotypes (Fig. 4B–D). A reduced number of dopaminergic neurons in borcs5-ko larvae was observed at 3 dpf compared to controls (Supplementary Fig. 2A, 2B). Hematoxylin and eosin staining revealed a reduction in overall brain size in borcs5-ko larvae compared to WT at 3 and 5 dpf (Fig. 4E, 4F; Supplementary Fig. 2G, 2H), suggesting a critical role for borcs5 in normal zebrafish brain development. Interestingly, we also observed larger ventricles in 5 dpf borcs5-ko brain compared to WT controls (Supplementary Fig. 2G).

**Figure 4:**
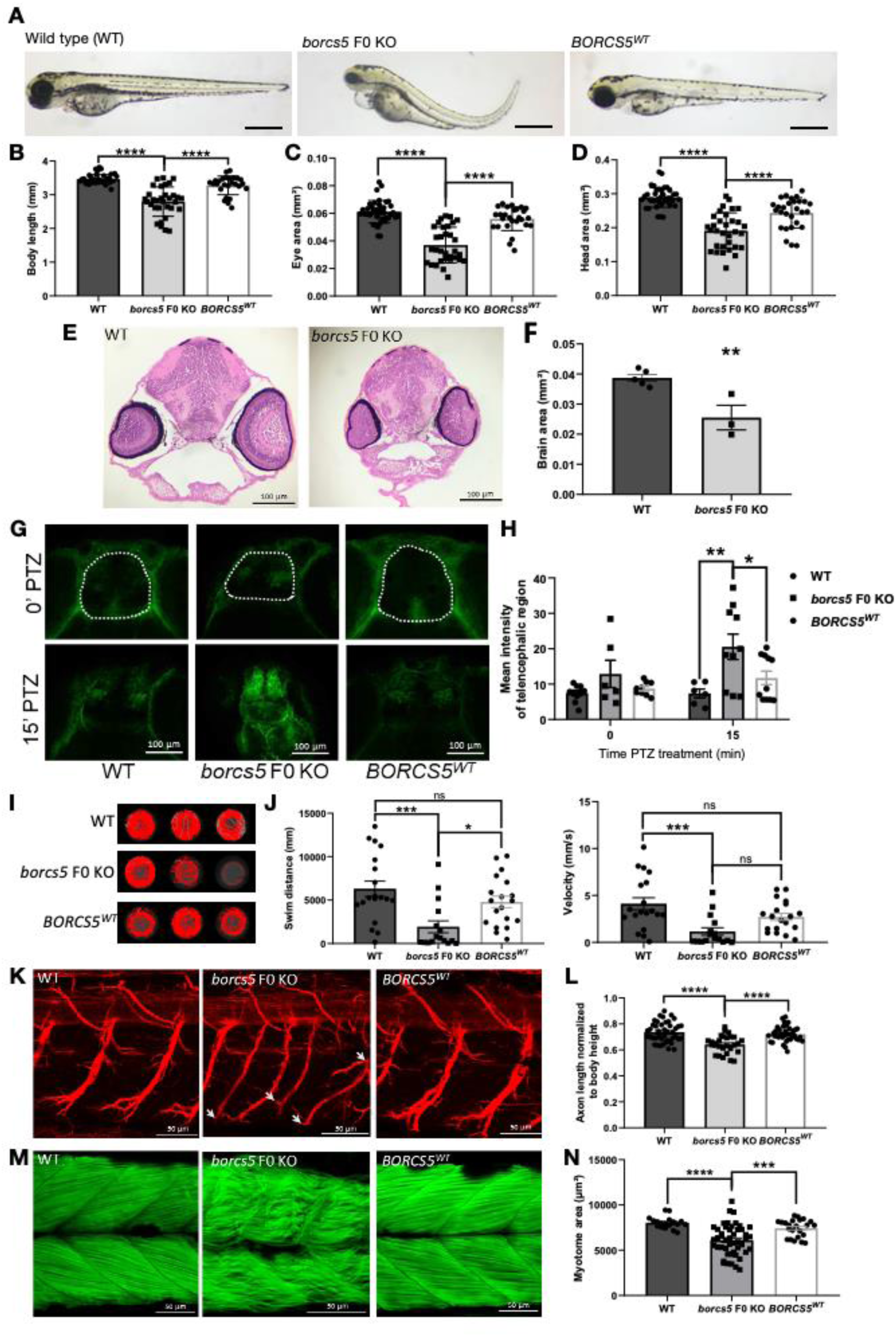
Zebrafish *borcs5* F0 KO larvae exhibit developmental defects and recapitulate patient symptoms. A. Morphology of zebrafish wild-type (WT), borcs5*-*ko and larvae injected with human *BORCS5* mRNA (*BORCS5^WT^*) at 3 days post-fertilization (dpf). Scale bars: 500 µm. B-D. Phenotypes observed for WT, borcs5*-*ko and *BORCS5^WT^*. Body length (B), eye size (C), and head size (D) of WT (n=40), borcs5*-*ko (n=32) and *BORCS5^WT^*(n=26-28) larvae at 3 dpf. E. H&E staining of midbrain sections of 3 dpf WT and borcs5*-*ko larvae. Scale bar: 100 μm. F. Quantification of brain area of WT (3 dpf, N=5) and borcs5*-*ko larvae (3 dpf, N=3). G, H. Neuronal activity induced by PTZ treatment (3 mM; 0, 15 min) in 4-dpf larvae was analyzed by quantification of mean intensity fluorescence of p-MAPK/ERK staining (surround in white). A significant increase in borcs5*-*ko (n=6-10) larvae compared to WT (n=6-10) and *BORCS5^WT^* (n=9-11) at 15 min treatment of PTZ was observed. Scale bars: 100 μm I. Representative swim trajectories of WT, borcs5*-*ko and *BORCS5^WT^* larvae at 5 dpf. (J) Quantification of swim distance and velocity in WT (n=19), borcks5-ko (n=16) and *BORCS5^WT^*(n=19). K. Acetyl tubulin staining of primary motor axon in WT, borcs5-ko and *BORCS5^WT^* larvae at 3 dpf. Scale bars: 50 μm. Defects in axon branching in borcs5-ko larvae are indicated by white arrows. L. Axon length normalized by body height in WT (n=10), borcs5-ko (n=6) and *BORCS5^WT^* (n=10). M. Muscle fibers visualized by phalloidin staining at 3 dpf. Scale bars: 50 μm. N. Quantification of dorsal or ventral myotome area between WT (n=9), borcs5-ko (n=10) and *BORCS5^WT^* larvae (n=12). All data are represented as the mean ± SEM. Statistical significance was calculated by One-way ANOVA followed by Tukey’s multiple comparisons tests, or Student’s T-test. *P < 0.05; **P < 0.01; ***P < 0.001; ****P < 0.0001; ns = not significant. n represents number of fish; N represents number of experimental repeats.

We next examined seizure susceptibility upon administration of pro-convulsive GABA receptor antagonist PTZ (3 mM) to 4 dpf larvae. Borcs5-ko larvae exhibited increased p-MAPK/ERK staining compared to WT at 15 min following PTZ treatment, which was rescued in *BORCS5*^WT^ (Fig. 4G, 4H), suggesting abnormal neuronal hyperactivation in borcs5-ko larval brain.

A marked decrease in locomotor activity was observed (i.e. free swimming) in borcs5-ko larvae at 5 dpf, as compared to WT and to *BORCS5^WT^* (Fig. 4I, J). We also found that borcs5-ko larvae had shorter and less branched axons in comparison to WT (Fig. 4K, L), and decreased area of dorsal and ventral myotomes with disorganization of somite structure and muscle fibers (Fig. 4M, N). The motor axonal and muscular defects in *borcs5* F0 KO zebrafish were rescued upon expression of the human *BORCS5* mRNA (*BORCS5*^WT^; Fig. 4K-N).

In addition to *BORCS5* LoF, we also functionally studied the missense variants R95Q and H99P via mRNA overexpression in borcs5-ko embryos. BORCS5^R95Q^ and BORCS5^H99P^ zebrafish displayed decreased body length, reduced head and eye size, albeit to a lesser extent than *borcs5* F0 KO zebrafish (Supplementary Fig. 2I-L). We also found reduced swimming distance, reflecting locomotor dysfunction, in BORCS5^R95Q^ and BORCS5^H99P^ larvae.

Taken together, these functional studies in zebrafish show that knock out of *borcs5* leads to neurodevelopmental defects, motor deficits and increased seizure susceptibility, indicating that *borcs5* plays a key role in the development and function of the CNS and supporting the pathogenic role of the identified variants.

### Effect of identified pathogenic variants on BORCS5 protein and BORC assembly

To analyze the effect of the individual coding variants on BORCS5 protein expression, BORCS5 WT and mutant constructs were transiently expressed in HEK293T. Cells expressing WT or the missense variants R95Q and H99P showed comparable BORCS5 protein expression. However, the LoF variants A106Pfs and Y139* exhibited shorter protein products and significantly lower protein levels compared to WT (Fig. 5A), confirming that these variants result in early truncation and reduced protein expression. Expression of L128Vfs showed multiple bands between 15 and 30 kDa (Fig. 5A), that likely are degradation products.

**Figure 5:**
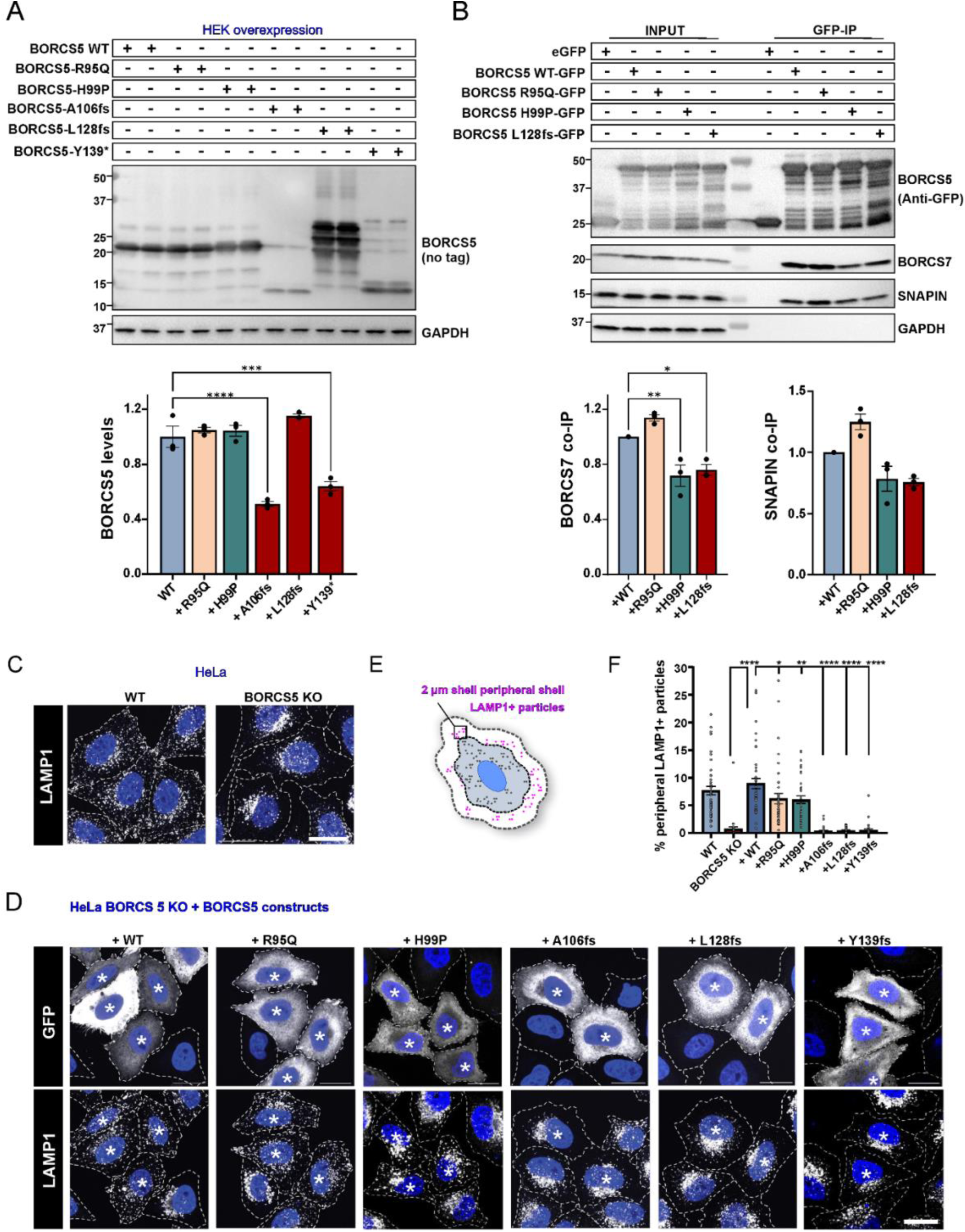
**Impact of BORCS5 variants on BORCS5 protein expression, BORC assembly and endolysosome distribution in cell lines**. A. Western blot analysis of BORCS5 protein in HEK293T cells transiently expressing the indicated BORCS5 WT or BORCS5 patient variants. GAPDH was used as a loading control. Quantification graph shows mean±SEM, N=3 independent experiments. Statistics: One way ANOVA with Dunnett’s post hoc (relative to WT construct), F_BORCS5_(5,12)=41.57, P<0.0001. ****p<0.0001, ***p=0.0002. B. Immunoprecipitation (IP) of GFP-tagged BORCS5 variants and their interaction (co-IP) with endogenous SNAPIN and BORCS7. GAPDH was used as a specificity control. Graph shows mean±SEM, N=3 independent experiments. Statistics: One way ANOVA with Dunnett’s post hoc (relative to WT construct), F_BORCS7_(3,8)=19.70, P=0.0005. **p=0.0055, *p=0.0136. F_SNAPIN_(3,8)=13.61, P=0.0017. C-D. Immunofluorescence microscopy shows endogenous LAMP1 (white puncta) distribution in untransfected WT and BORCS5 KO HeLa cells as a control. BORCS5-KO HeLa cells were transiently co-transfected with the indicated BORCS5 constructs and GFP. Immunofluorescence microscopy shows endogenous LAMP1 distribution in GFP+ transfected cells (indicated by asterisk). Nuclei were labeled with DAPI (blue), and cell edges were outlined by fluorescent phalloidin (indicated by dashed lines). Scale bars: 20 μm. E. Schematic depicts the analysis performed to quantify the percentage of LAMP1+ endolysosomes present in a 2-µm peripheral shell. F. Quantification shows mean±SEM. Statistics: One-way ANOVA with Dunnett multiple comparisons test (compared to WT construct), F_peripheral lysosomes_ (7,289)=30.96, P<0.0001, *p<0.05, **p=0.0098, ****p < 0.0001.

Furthermore, we assessed the impact of *BORCS5* pathogenic variants on the interaction with the BORC subunits SNAPIN and BORCS7 using GFP trap precipitation upon expression of C-terminally GFP-tagged BORCS5 constructs (Fig. 5B). We found 30% less endogenous BORCS7 protein co-immunoprecipitation with the H99P and L128Vfs constructs, whereas R95Q did not impact the BORCS5-BORCS7 interaction (Fig. 5B). In contrast, the interaction with SNAPIN was not significantly affected by the tested BORCS5 variants (Fig. 5B). The truncating variants A106Pfs and Y139* were not included in this experiment due to low baseline protein expression levels (Fig. 5A).

### BORCS5 LoF alleles, but not the missense variants R95Q and H99P, cause abnormal perinuclear clustering of lysosomes

Since the BORC complex regulates the anterograde transport of endolysosomes from the perinuclear region towards the cell periphery,^8,12^ the distribution of endolysosomes was analyzed in HeLa cells transiently overexpressing WT BORCS5 or BORCS5 with pathogenic variants (Fig. 5C-F). Loss of endogenous BORCS5 led to a perinuclear distribution of endolysosomes, reflected by a 90% reduction of LAMP1+ puncta in the cell periphery of BORCS5-KO cells, a phenotype that could be fully reversed by reintroducing BORCS5 WT (Fig. 5C, 5F). Expression of BORCS5 LoF variants A106Pfs, L128Vfs, and Y139* did not rescue the perinuclear distribution of endolysosomes, confirming that these variants completely disrupt BORCS5 function (Fig. 5D, F). In contrast, expression of the missense variants R95Q and H99P led to ∼30% fewer peripheral lysosomes compared to WT, suggesting a partial rescue of perinuclear endolysosomal clustering (Fig. 5D, 5F).

To examine BORC assembly and lysosomal distribution in patient derived cells, skin fibroblasts were cultured from patients carrying missense variants (F-I:1 and F-I:2 with compound heterozygous variants R95Q/L128Vfs and patient F-II:1 with homozygous H99P/H99P) and from a patient with a complete LoF variant (F-V:6 with homozygous Y139*/Y139*). All lines exhibited lower BORCS5 protein, with the two fibroblast lines R95Q/L128Vfs showing a 25-40% reduction, the H99P/H99P line with a 60% reduction and the Y139*/Y139* line having 93% lower BORCS5 protein as compared to control (Fig. 6A, B). Additionally, as it is known that the loss of one BORC subunit results in a reduced expression of other subunits,^17^ we examined the endogenous levels of BORC complex subunits SNAPIN and BORCS7. Fibroblasts from the two R95Q/L128Vfs patients showed 25-35% lower BORCS7, H99P/H99P showed 50% less and Y139*/Y139* fibroblasts had 75% lower protein (Fig. 6A, C). In contrast, SNAPIN was exclusively reduced in fibroblasts from the Y139*/Y139* patient, but not in the other lines (Fig. 6A, D).

**Figure 6:**
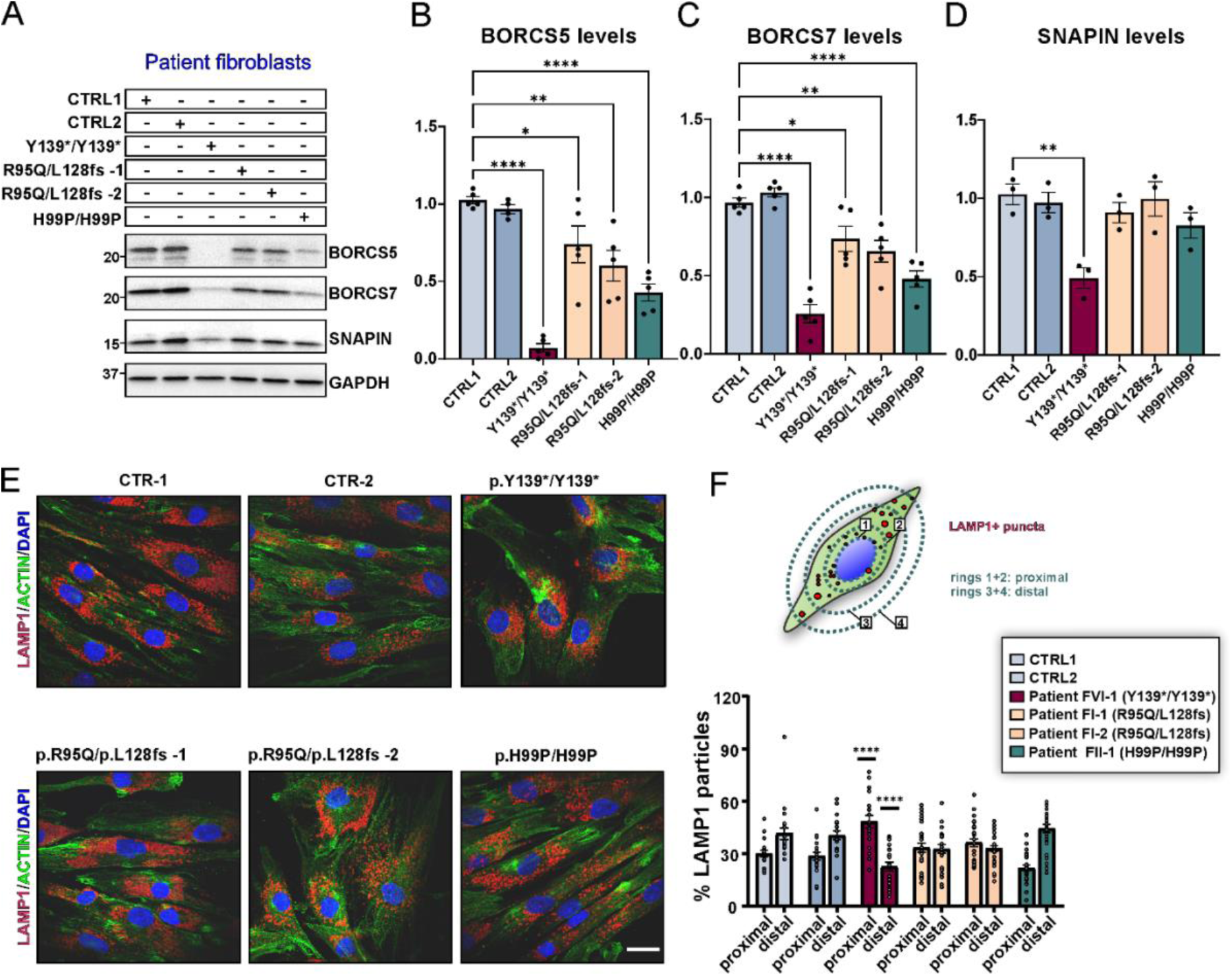
**BORC-related protein expression and endolysosomal distribution in BORCS5 patient fibroblasts**. A. The western blot shows relative levels of BORCS5, SNAPIN and BORCS7 in patient fibroblasts compared to two independent control lines. GAPDH was used as a loading control. B-D. Graphs represent mean±SEM, N=3-5 independent experiments. Statistics: One way ANOVA with Dunnett’s post hoc (compared to CTRL1), FOR CS5(5,23)=24.56, P<0.0001; F_BORCS7_(5,24)=27.02, P<0.0001; F_SNAPIN_(5,12)=6.704, P=0.0034, *p<0.03, **p<0.003, ****p<0.0001. E. Immunofluorescence microscopy shows endogenous LAMP1 puncta distribution in control or BORCS5 patient fibroblasts, quantified according to the schematic F. Concentric rings of 1.5 increment were designed using Fiji, using individual nuclei as reference. Puncta within rings 1 and 2, were designated proximal whereas those in rings 3 and 4 were considered distal to the nucleus. Graph represents mean±SEM from N=3 independent experiments. Statistics: Two-way ANOVA F_Interaction_(5,276)=22.86, P<0.0001, with Šidák post hoc (compared to CTRL and CTRL2).

To evaluate endolysosomal distribution in BORCS5 mutant patient fibroblasts, we compared the percentage of LAMP1+ puncta located proximally vs distally to the nucleus (Fig. 6E, F). BORCS5 Y139*/Y139* patient fibroblasts had 60% more endolysosomes distributed proximal to the nucleus and 40% fewer endolysosomes distributed towards the cell periphery (Fig. 6E, F). However, no changes in endolysosomal distribution were found in BORCS5 R95Q/L128Vfs and H99P/H99P patient fibroblasts (Fig. 6E, F).

Altogether, these results indicate that the A106Pfs, Y139*, and L128Vfs are complete LoF alleles, while the H99P is a hypomorphic allele associated with reduced endogenous protein levels. Complete loss of BORCS5 likely affects the assembly of BORC and the protein levels of its subunits, whereas the H99P allele selectively affects the interaction with BORCS7. The R95Q allele is normally expressed and is not likely to affect the interaction with other BORC subunits and BORC assembly. Furthermore, BORCS5 complete LoF variants lead to perinuclear lysosomal clustering, whereas the anterograde transport of endolysosomes is at least in part preserved in the presence of the missense BORCS5 variants R95Q and H99P.

### Both LoF and missense BORCS5 variants lead to lysosomal dysfunction

Loss of BORC subunits, including BORCS5, has been shown to reduce autophagosome-to-lysosome fusion^15^, leading to intracellular accumulation of autophagosomes. Consistent with this notion, *BORCS5* knockout in HeLa led to a significant increase in the number of puncta positive for the autophagosomal marker LC3 (LC3+ puncta), a phenotype that could be restored by expressing WT BORCS5 (supplementary fig. 3A-C). While the expression of all LoF variants failed to rescue the KO phenotype, expression of the missense variants R95Q and H99P restored the LC3+ puncta to WT levels (supplementary fig. 3A-C), suggesting that these variants do not cause impairment in autophagosome clearance.

We further investigated endolysosomal function in BORCS5 patient fibroblasts. Transmission electron microscopy (TEM) revealed the presence of multilamellar bodies, structures related to dysfunctional lysosomes,^20^ in fibroblasts with complete BORCS5 LoF (Y139*/Y139*) as well as in those carrying the missense variant R95Q (R95Q/L128Vfs) (Fig. 7A, B). No differences were found in the number of autophagic structures or lysosomes between controls and fibroblasts with LoF or missense variants (supplementary fig. 4A).

**Figure 7:**
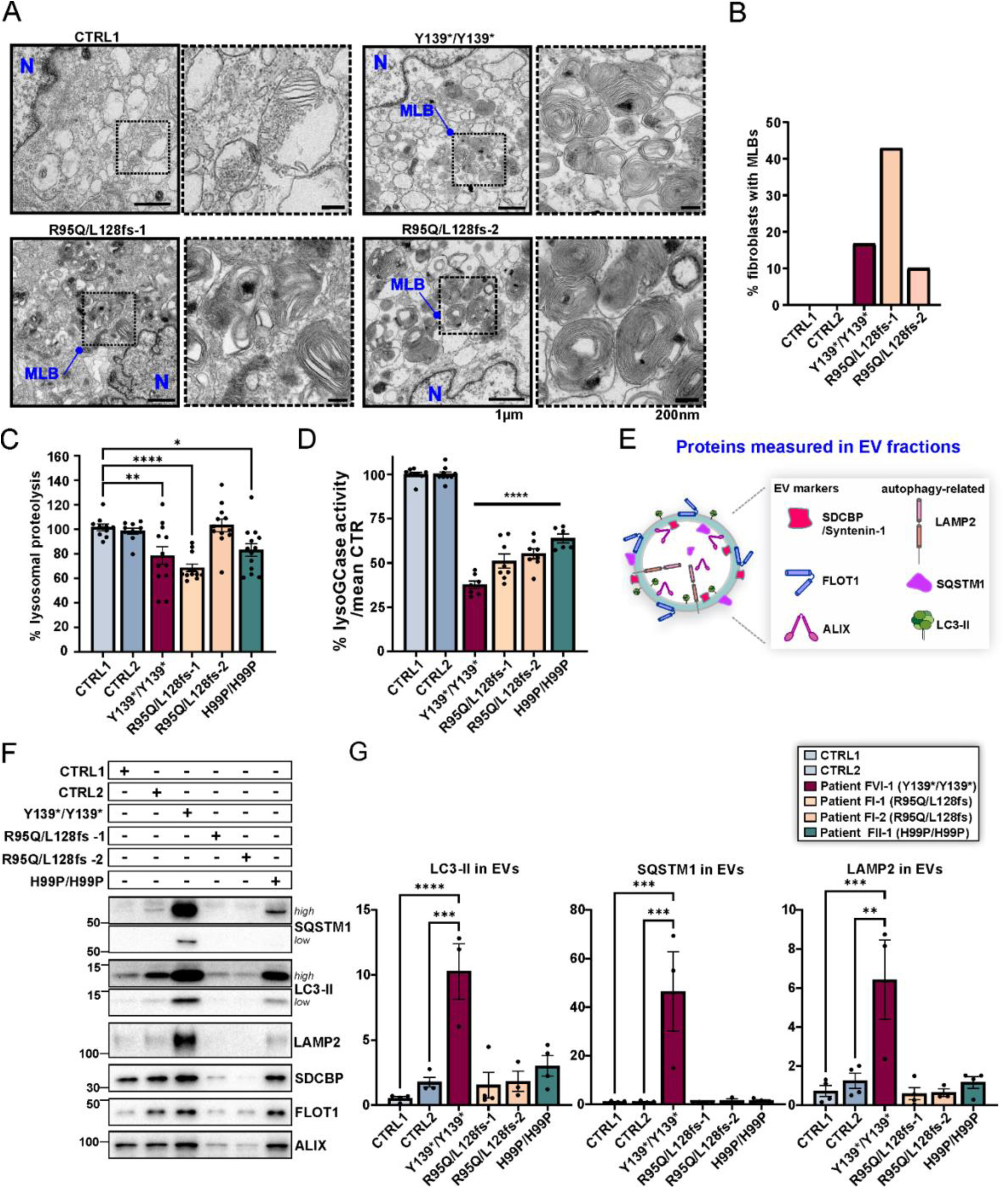
Lysosomal dysfunction in *BORCS5* patient fibroblast lines. A. TEM of fibroblasts from the indicated *BORCS5* genotypes. Outlined insets are presented at higher magnification on the right, indicated by dashed lines. Abbreviations: N: Nucleus; MLB: Multilamellar body. B. Graph represents the percentage of fibroblasts exhibiting MLBs under TEM with N=9 to 18 individual cells visualized per fibroblast line. C. The efficiency of lysosomal proteolysis was assessed upon administration of 25µg/mL of DQ Green or Red BSA for 5 h, in 10,000 single cell events, via flow cytometry. Graph represents the mean±SEM of the fold change in median fluorescence intensity, normalized to the mean of control lines. CTR-1 and CTR-2: N=9; p.Y139*/p.Y139* and p.R95Q/L128Vfs*86: N=8; p.H99P/H99P: N=9 independent experiments. Statistics: One way ANOVA with Dunnett’s post hoc (compared to CTRL1). Proteolysis(5,63)=8.687, P<0.0001. ** p=0.007, **** p<0.0001, * p=0.038. D. Lysosomal GCase in control or patient fibroblasts was assessed upon administration of 250µM PFB-FDGlu for 30min, in 10,000 single cell events, via flow cytometry. Graph represents the mean±SEM of the % change in median fluorescence intensity, normalized to the mean of control lines. CTR-1 and CTR-2: N=7; p.Y139*/p.Y139*: N=4, p.R95Q/L128Vfs*86 N=4; p.H99P/H99P: N=3 independent experiments. Statistics: One way ANOVA with Dunnett’s post hoc (compared to CTRL1). F_lysoGCase_(5,41)=121.3, P<0.0001. **** p<0.0001. E. Summary of proteins analyzed in exosome/extracellular vesicle (EV) fractions isolated from identical volumes of fibroblast conditioned medium via ultracentrifugation. F-G. WB of EVs and quantification of autophagy/lysosome-related markers normalized to controls. Graphs show mean±SEM, N=3-4 independent experiments. Statistics: One way ANOVA with Tukey’s multiple comparisons test (mean of each column compared to the mean of every other column), F_LC3-II_(5,16)=12.51, P<0.0001. **** p<0.0001, *** p=0.0002; SQSTM1(5,16)=10.65, P=0.0001. *** p=0.0002; F_LAMP2_(5,16)=8.572, P=0.0004. *** p=0.0007, ** p=0.0017.

The efficiency of lysosomal proteolysis was measured by flow cytometry of single-cell fluorescence intensity in fibroblasts upon 5 h of DQ BSA hydrolysis (Fig. 7C). Proteolysis was reduced by ∼20% in all fibroblast lines with both LoF and missense pathogenic variants as compared to controls (Fig. 7C). Examination of total cell lysates by western blotting showed comparable amounts of the lysosomal membrane protein LAMP2 that together with the TEM data confirmed that deficient lysosomal proteolysis was not due to a lower number of lysosomes (supplementary fig. 4B). Notably, the activity of lysosomal hydrolase glucocerebrosidase (GCase) was ∼40% lower in cells with BORSC5 missense variants and ∼60% lower in the LoF line (Fig. 7D), whereas GCase protein and its lysosomal transporter LIMP2 were not changed (supplementary fig. 4B-D). Thus, both BORCS5 LoF and missense variants lead to decreased lysosomal GCase activity and impaired general lysosomal proteolysis.

Since loss of BORCS5 function results in lysosomal dysfunction (Fig. 7A-D), we hypothesized that the release of exosomes/extracellular vesicles (EVs) may be altered under these conditions.^16,21,22^ EV fractions isolated from identical volumes of conditioned medium across all fibroblast lines were positive for standard exosomal markers^23^ Syntenin-1 (SDCBP), flotillin-1 (FLOT-1) and ALIX (supplementary fig. 4F).

Importantly, EVs from both the BORCS5 complete LoF (Y139*/Y139*) and the missense fibroblast line (H99P/H99P) showed increased levels of the autophagy markers LC3-II and p62/SQSTM1, and endolysosomal membrane marker LAMP2 (Fig. 7E-G), indicating that BORCS5 LoF leads to lysosomal dysfunction and consequent release of undegraded cargo via EVs.

### Defects in endolysosome distribution and lysosomal function in BORCS5-mutant patient iPSC-derived forebrain neurons

Given that all *BORCS5*-mutant patients exhibited exclusively neurological symptoms, we further examined the impact of complete BORCS5 LoF and the R95Q missense variant on trafficking and endolysosomal function in a human neuronal model.

Patient fibroblasts from the two affected subjects from family F-1 carrying the compound heterozygous BORCS5 variants R95Q/L128Vfs were reprogrammed into induced pluripotent stem cells (iPSC). In addition, a control iPSC line was used to generate an isogenic CRISPR/Cas9 *BORCS5* knockout line (BORCS5-KO). All iPSC lines were differentiated into human forebrain neurons via Ngn2 overexpression.^24^

Using the fluorescent lysosomotropic dye Lysotracker Red, we found that BORCS5-KO neurites contained 60% fewer endolysosomes as compared to isogenic control neurons (Fig. 8A, B), in agreement with previous reports.^11,12,14^ However, no changes in endolysosomal distribution were observed in neurites of BORCS5-R95Q/L128Vfs versus WT neurons (Fig. 8A, B).

**Figure 8:**
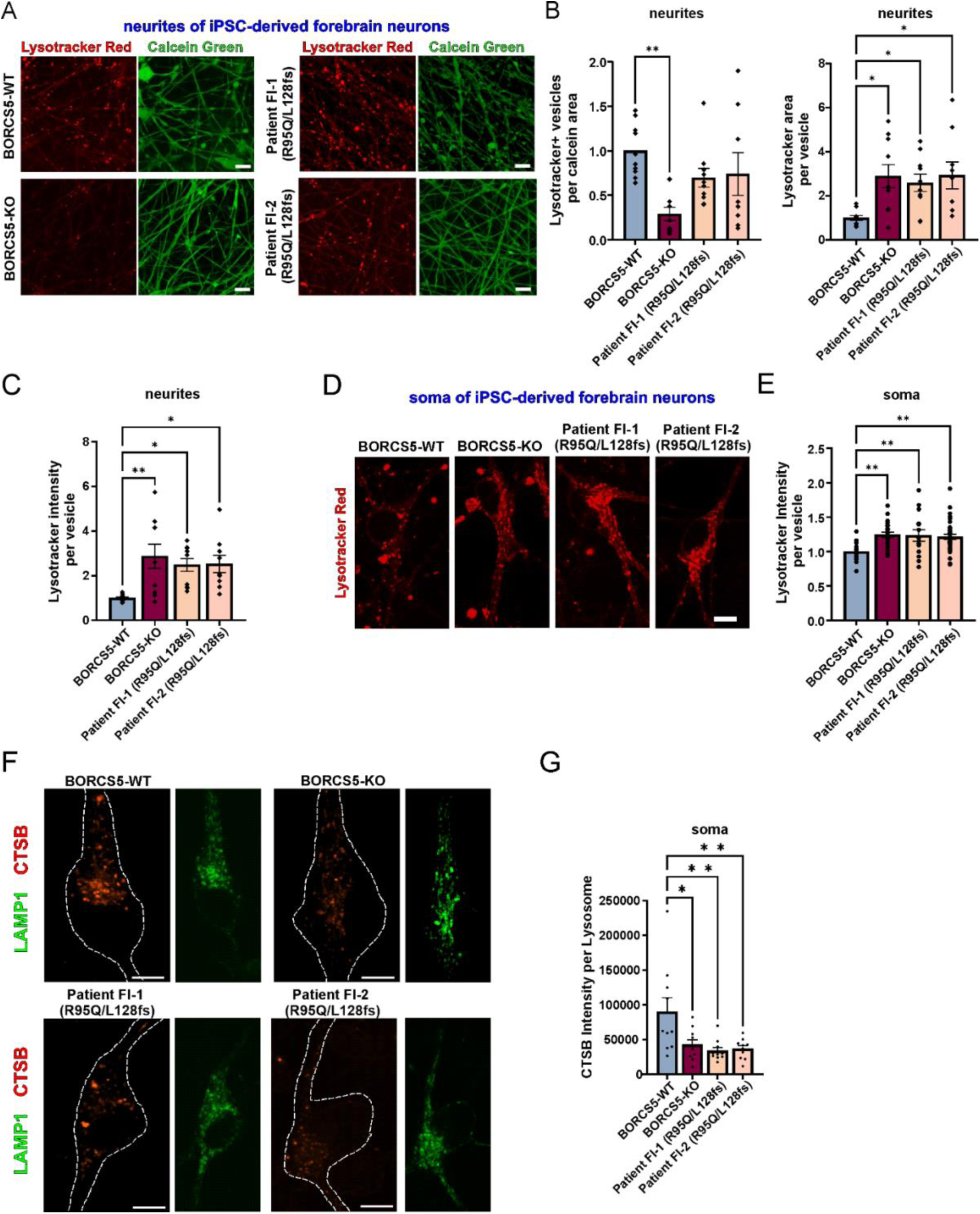
Lysosome-dispersal activity of patient-specific *BORCS5* variants. Increased LysoTracker and decreased lysosomal enzyme activity in neurons with patient-specific *BORCS5* variants. A. Imaging of Lysotracker Red endolysosomes (red) in neurites of live iPSC-derived forebrain neurons (labeled with calcein, green) from WT, isogenic BORCS5-KO or R95Q/L128fs patient iPSC lines. B, C Graphs show mean±SEM, N=3 independent experiments. Statistics: One way ANOVA with Dunnett’s post-hoc. F_vesicle number_ (3,34)=5.250, P=0.0044; F_vesicle area_(3,35)=5.373, P=0.0038; F_signal intensity_(3,35)=4.362, P=0.0104. D. Imaging of Lysotracker Red endolysosomes (red) in the soma of iPSC-derived forebrain neurons from the indicated lines. D. Graph shows mean±SEM, N=3 independent experiments. Statistics: One way ANOVA with Dunnett’s post-hoc. F_signal intensity_(3,87)=5.624, P=0.0014. F. Fluorescence microscopy examination of LAMP1+ stained endolysosomes (green) and cathepsin B (CTSB) activity-derived fluorescent signal (red) in the soma of iPSC-derived forebrain neurons from the indicated lines. G. Graph shows mean±SEM, N=3 independent experiments. Statistics: One way ANOVA with Dunnett’s post-hoc. F_CTSB activity soma_(3,36)=5.478, P=0.00333. Dots represent individual cells.

Focusing on endolysosomal function, we found that compared to WT both BORCS5-KO and R95Q/L128Vfs neurons, exhibited a 3-fold higher Lysotracker Red area as well as intensity per vesicle (Fig. 8A-C). In addition, Lysotracker Red intensity per vesicle was 25% higher in the soma of both BORCS5-KO and BORCS5-R95Q/L128Vfs patient neurons (Fig. 8D-E). To further address lysosomal activity within these enlarged vesicles, we used the Magic Red cathepsin B substrate that emits fluorescence upon cleavage by active cathepsin B in neurons (Fig. 8F, G). This assay showed 50% lower cathepsin B activity within endolysosomes of the soma of BORCS5-KO and BORCS5-R95Q/L128Vfs neurons as compared to WT neurons (Fig. 8F, G).

These findings support the notion that only complete BORCS5 LoF disrupts anterograde axonal transport of endolysosomes, while neurons expressing the R95Q missense variant retain this function. However, all *BORCS5*-mutant neurons exhibited enlarged and functionally impaired lysosomes, suggesting that both complete LoF and pathogenic missense variants contribute to neuronal lysosomal dysfunction.

## Discussion

We provide genetic and functional evidence establishing *BORCS5* as a human disease-associated gene responsible for a severe neurological disorder characterized by a broad spectrum of phenotypes, from profound neurodevelopmental defects to infantile-onset neurodegeneration. Furthermore, we uncovered a novel role for BORCS5 in the regulation of lysosomal function, expanding beyond its well-established function in mediating anterograde lysosomal trafficking.

*BORCS5* complete LoF variants were clinically associated with a severe, perinatally lethal phenotype with neuropathological evidence of NAD, a condition associated with extreme and rapidly progressive neurodegeneration. Similar pathological findings have also been reported in murine models of Borcs5 and Borcs7 complete LoF.^11,13^

These fetuses also displayed a severe developmental phenotype, characterized by agenesis of the corpus callosum, small brainstem and cerebellum, and aqueductal stenosis with supratentorial hydrocephalus. This suggests a critical role for BORCS5 in the development of the corpus callosum and midbrain-hindbrain structures, similarly to what has been shown in cases with pathogenic variants in *KIF5C and KIF2A*, which encode other critical components of the organelle anterograde transport machinery.^25,26^

Conversely, carriers of missense variants exhibited a chronic neurodegenerative presentation with infantile onset, associated with progressive movement disorders and severe spasticity. Neuroradiological findings in the latter group showed a profound neurodegenerative phenotype with early-onset global cerebral atrophy and hypomyelination, closely resembling features observed in subjects harboring *BORCS8* variants.^17^

The *borcs5* F0 knockout zebrafish model recapitulated many of the phenotypic features observed in *BORCS5*-mutant cases, including microcephaly, ventriculomegaly, movement disorders and epilepsy. Thus, the *borcs5-*ko zebrafish model may be valuable to shed more light on the underlying pathophysiological mechanisms of the neurological defects associated with bi-allelic *BORCS5* variants.

Our study reveals a novel role for BORCS5 as a regulator of lysosomal function, expanding on its recognized role in lysosomal trafficking. Specifically, all patient fibroblast lines showed decreased lysosomal degradation efficiency and reduced lysosomal GCase activity compared to controls. BORCS5-KO and R95Q/L128Vfs neurons exhibited enlarged endolysosomes and reduced activity of lysosomal enzymes, consistent with endolysosomal dysfunction.^27^ Moreover, the protein profile of EVs released from BORCS5 LoF fibroblasts indicated increased release of undigested autophagic and lysosomal protein cargos due to reduced endolysosomal degradation.^16,21,22^,

Consistent with this notion was the presence of multilamellar bodies in BORCS5 patient fibroblasts since similar structures have been described in patients with lysosomal storage disorders, including Niemann Pick disease types A and C, as well as PD cases carrying pathogenic *GBA1* variants.^20,28–30^ Moreover, subjects carrying *BORCS5* missense variants exhibited clinical and radiological features resembling those seen in infantile-onset lysosomal storage disorders, such as gangliosidosis, fucosidosis, and neuronal ceroid lipofuscinoses.^31^

The underlying mechanism of lysosomal dysfunction caused by *BORCS5* pathogenic variants remains to be fully elucidated. We found reduced cathepsin B activity, which in turn may impair lysosomal GCase activity through deficient processing of prosaposin into the endogenous GCase activator saposin C.^32^ Cathepsin B is also involved in processing of progranulin to granulin fragments that can modulate both GCase and lysosomal function.^33,34^ Additionally, BORCS5 deficiency may disrupt lysosomal maturation,^16^ leading to the accumulation of functionally immature late endosomes or impair the intracellular trafficking of hydrolases from the endoplasmic reticulum/trans-Golgi network to lysosomes, resulting in reduced lysosomal hydrolase activity.

The role of BORCS5 in lysosomal function may have important implications for other neurological disorders characterized by lysosomal dysfunction, including PD and dystonia. Intriguingly, several elements link BORCS5 dysfunction to PD pathogenesis. The genes encoding Cathepsin B and glucocerebrosidase, whose activity is reduced by *BORCS5* mutations, are established risk factors for PD.^35–38^Additionally, we observed that the axonal spheroids in one of the *BORCS5*-mutant cases (F-VII:1) with NAD displayed strong staining for α-synuclein, the most abundant protein found in Lewy bodies (LBs). Moreover, the most common genetic cause of NAD are mutations in *PLA2G6*, a gene associated with a broad spectrum of neurodegenerative conditions, spanning from infantile NAD to early-onset PD^39,40^ and pathologically characterized by diffuse and severe α-synuclein pathological aggregation and formation of LBs.^41,42^ While we did not observe LBs in the brain of *BORCS5*-mutant cases, this was likely due to their prenatal lethality. Finally, we observed loss of dopaminergic neurons in *borcs5* KO zebrafish, a result which corroborates the response to dopaminergic treatment of the movement disorders in some of the *BORCS5*-mutant cases.

Pathogenic mutations in several components of the HOPS complex, including *VPS11*, *VPS16,* and *VPS41* that is known to functionally interact with BORC and is involved in autophagosome-lysosome fusion and late-endosome to lysosome maturation,^15,16^ have been described to be causative for cases with movement disorders like ataxia and dystonia.^3,43–47^ Thus, lysosomal dysfunction underlying these genetic defects might be a unifying factor in the pathogenesis of these movement disorders.

In conclusion, through mechanistic investigations of *BORCS5* disease-causing variants, we uncovered a novel role for BORCS5 in the regulation of lysosomal function, in addition to its well-established role in anterograde lysosomal movement. This dual function may explain the distinct clinical phenotypes observed in cases with different *BORCS5* mutations. Complete BORCS5 LoF leads to severe neurodevelopmental defects and diffuse prenatal NAD, whereas missense mutations impair lysosomal activity without disrupting lysosomal distribution, resulting in infantile-onset neurodegeneration. These insights provide a foundation for further research into BORC-mediated lysosomal homeostasis, with potential therapeutic implications for neurodegenerative disorders.

## Material and methods

### Patient ascertainment and clinical and molecular studies

Individuals and/or their legal guardians recruited for this study gave informed consent for their participation. This study received approval from the Review Boards and Bioethics Committees at University College London Hospital (project 06/N076). Informed consent was obtained for all individuals enrolled in the study, including permission to publish de-identified clinical images and videos where available.

Comprehensive clinical data were collected from all affected individuals, including detailed phenotypic features, family history, photographs, videos, clinical notes, and brain MRI findings. All brain MRIs were reviewed and interpreted by an experienced pediatric neuroradiologist.

WES and Sanger sequencing were performed independently in different research and clinical laboratories using established protocols.^48–50^ In the index family (F-I), WES was carried out on the two affected siblings and both parents. Based on the consanguineous family structure, analysis focused on identifying rare bi-allelic coding and essential splice-site variants. *BORCS5* emerged as the sole candidate gene from this analysis.

To identify additional families with bi-allelic *BORCS5* variants, we adopted a genotype-first approach. Systematic re-analysis and screening of large-scale sequencing datasets were performed across multiple collaborative research networks and data-sharing platforms. These included GeneMatcher (Families F-II, F-IV, F-V), Igenomix (Family F-VI), CENTOGENE (Family F-III), GeneDx (Family F-VII), as well as screening of datasets from the UCL Queen Square Genomics platform, the 100,000 Genomes Project, Solve-RD, Baylor Genetics, Lifera Omics, Genesis, ClinVar, VarSome, and several smaller local and private diagnostic or research laboratories.

Allele frequencies of the identified *BORCS5* variants were evaluated in population databases, including gnomAD v4.1.0 (covering ∼800,000 individuals, ∼5% of whom are of South Asian descent), the UCL Queen Square Genomics Database (∼35,000 individuals, enriched for underrepresented populations), and the Igenomix internal database (∼65,000 individuals, ∼20% of whom are of Arab ancestry).

Skin fibroblasts were obtained from affected subjects from F-I (two lines), F-II (one line), and F-V (one line). Two unrelated fibroblast lines from gender and age-matched control subjects were also included in the study.

### Culture and transfection of human cell lines

Cells were maintained at 37°C in a 5% CO_2_ incubator and routinely tested for mycoplasma contamination using PCR-based detection (Venor GeM Mycoplasma Detection Kit (Sigma, MP0025). HEK-293 FT and HeLa cells were cultured in Dulbecco’s modified Eagle’s Medium (DMEM; Gibco, 11995-065) supplemented with 10% heat-inactivated fetal bovine serum (HI-FBS; Benchmark, 100-106). Fibroblasts were cultured in Dulbecco’s modified Eagle’s Medium (DMEM; Gibco, 11995-065) supplemented with 15% heat-inactivated fetal bovine serum (HI-FBS; Benchmark, 100-106), 10 U/ml penicillin and 10 μg/ml streptomycin (1% Pen Strep; Gibco, 15140-122). Cells were passaged with trypsin (TrypLE; Invitrogen, 12605-010) for maintenance.

HeLa and HEK-293T cells were transiently transfected with 0.8 μg plasmid DNA using 2 μl Lipofectamine 2000 (Invitrogen), according to the manufacturer’s instructions. Approximately 24 h after transfection, HEK-293T cells were harvested and HeLa cells were replated onto 12-mm coverslips coated with collagen. HeLa cells were then cultured for an additional 24 h before fixation and immunofluorescence labelling.

### HeLa BORCS5 CRISPR/Cas9 knockout generation

Briefly, two 20-base pair (bp) targeting sequences (GCTCAACAGCATGCTGCCCG and AGCAGATCCAGAAAGTGAAC) were synthesized (Eurofins) and introduced separately into the px330 plasmid (Addgene). HeLa cells were co-transfected with both plasmids and re-seeded after 72 h to allow single colony formation. After 12 days, genomic DNA was extracted from individual colonies, and cleavage of the target sequence was tested by PCR using a pair of primers (ATCTGCGGGACTGTGTCCCT and CAGATTTTCATGCCAGCCGG), which produced a 99-bp smaller band in KO cells relative to WT cells. The KO was confirmed by Sanger sequencing and immunoblotting.

### Protein extraction and western blot analysis

HEK-293T and fibroblasts were washed with PBS and protein was extracted in RIPA buffer (Boston BioProducts, BP-115-5x; 50 mM Tris-HCl, pH 7.4, 150 mM NaCl, 1% Nonidet P-40 substitute, 0.5% Na-Deoxycholate, 0.1% sodium dodecyl sulfate), supplemented with protease (Roche, #11836170001) inhibitors. Depending on the protein marker of interest, 20 to 50 µg of total protein were electrophoresed using Tris-Glycine gradient gels (Novex™ 4-12% Tris-Glycine Mini Gels, WedgeWell™ format, 12-well, XP04122BOX; 15-well-WedgeWell, XP04125BOX; Novex™ WedgeWell™ 4-20% Tris-Glycine Mini Gels, 12-well, XP04202BOX; 15-well-WedgeWell, XP04205BOX, Thermo Fisher Scientific), semi-dry transferred onto PVDF membranes (Trans-Blot Turbo System, 1704275, BioRad) and incubated for 1 h at room temperature with blocking buffer (TBS-T; Tris-buffered saline with 0.1% Tween-20 (Applichem, A1389,0500)) supplemented with 1% w/v bovine serum albumin (Sigma-Aldrich, A9647)). The primary antibody was diluted in a blocking agent and incubated overnight at 4°C with mild shaking, followed by TBS-T washes, and incubation with appropriate secondary antibodies (also diluted in blocking buffer). Chemiluminescence was visualized and analyzed using the Chemidoc Software (BioRad).

### Immunoprecipitation

For GFP precipitation, cells were transfected with cDNA of wild-type or mutant BORCS5-GFP, using GFP alone as control. After 24 hours of expression, cells were harvested and subsequently lysed by sonication in 10mM Tris-CL pH 7.5, 150 mM NaCl, 0.5 mM EDTA, 1% Triton X 100, 10%glycerol) supplemented with protease inhibitor cocktail (Roche). Cell debris was removed by centrifugation and the supernatant was subjected to immunoprecipitation. A fraction of the supernatant was kept as an input sample. ChromoTek GFP-Trap Agarose beads (Proteintech, USA) blocked with 5% BSA in lysis buffer were incubated with residual supernatant for 2 hours at 4°C. Beads were collected by centrifugation at 800xg and 4°C and washed five times with 500 μl lysis buffer. Precipitates were eluted with Laemmli buffer (Bio-Rad) containing β-mercaptoethanol at 55°C for 30 min. The immunoprecipitated samples and inputs were analyzed by immunoblotting.

### Lysosomal distribution and LC3 levels analysis by imaging

Quantification of LAMP1 distribution in HeLa cells was performed as previously described (Williamson et al., 2022 PMID: 35819772). Briefly HeLa cells grown on 6-well plates were transiently transfected with 0.8 μg of each plasmid DNA using 2 μl Lipofectamine 2000 (Invitrogen), according to the manufacturer’s instructions. Approximately 24 h after transfection, cells were replated on collagen-coated coverslips in 24-well plates at 40,000 cells per well. Cells were then cultured for an additional 24 h to allow rescue of the BORC phenotype. Cells were then fixed in 4% w/v paraformaldehyde (Electron Microscopy Sciences) in PBS for 20 min, permeabilized and blocked with 0.1% w/v saponin, 1% w/v BSA (Gold Bio) in PBS for 20 min, and sequentially incubated with primary and secondary antibodies (mouse anti-LAMP1 (H4A3, DSHB, IF 1:500), chicken anti-GFP (A10262, Thermo Fisher, IF 1:500), Alexa Fluor 555-conjugated donkey anti-mouse IgG (A-31570, Thermo Scientific, IF 1:1,000), Alexa Fluor 488-conjugated Goat anti-Chicken IgY (H+L) (A-11039, Thermo Scientific, IF 1:1,000), Alexa FluorTM 647-conjugated phalloidin (A22287, Thermo Scientific, IF 1:50) diluted in 0.1% w/v saponin, 1% w/v BSA in PBS for 30 min at 37°C. Coverslips were washed three times in PBS and mounted on glass slides using Fluoromount-G (Electron Microscopy Sciences) with DAPI. Z-stack cell images were acquired on a Zeiss LSM 900 inverted confocal microscope (Carl Zeiss) using a Plan-Apochromat 63X objective (NA=1.4). Maximum intensity projections and final composite images were created using ImageJ/Fiji (https://fiji.sc/). The final images were subjected to shell analysis (as shown in the schematic of Fig. 5C). Briefly, cells exhibiting morphologies where perinuclear clusters of lysosomes were situated too close to the plasma membrane were excluded from analysis. Cell outlines were traced in Fiji (https://imagej.net/software/fiji/) using the phalloidin staining as track, and the total fluorescence of LAMP1 signal was measured. The cell outline was then shrunk by 2 μm using the “enlarge” function in Fiji. The LAMP1 signal intensity was measured in this smaller shell and subtracted from the larger value. The intensity of LAMP1 signal within the peripheral 2-μm shell was then plotted as percentage of total cellular LAMP1 signal. For statistical analysis, we performed a one-way analysis of variance (ANOVA), followed by multiple comparisons using Dunnett’s test. All statistical analyses were conducted using Prism version 9 (GraphPad Software). For the LC3B quantification cells were plated, stained (rabbit anti-LC3 (3868, Cell Signaling, IF 1:200), Alexa Fluor 555-conjugated goat anti-rabbit IgG (A-21428, Thermo Scientific, IF 1:1,000)), and images were acquired as described earlier in this paragraph. LC3B particles were counted for each cell using the “Analyze particles” function of Fiji.

For fibroblasts, lysosomal distribution in confocal images was analyzed by ImageJ/Fiji, with modifications in a published protocol.^51^ Specifically, fibroblast coverslips were co-stained for LAMP1, actin to outline the cell area, and DAPI to define nuclei. Four different regions per coverslip were imaged by confocal microscopy. In each image, non-overlapping individual fibroblasts were analyzed using the DAPI channel as reference to define four regions of interest (ROI) per fibroblast, as shown in the schematic of Fig. 5F. The first ROI was the outline of the nucleus (oval-shaped ring number 1) and an additional three concentric rings of 1.5 increments towards the cell periphery (oval-shaped rings 2 to 4) were designed. Next, using the LAMP1 channel (type 8-bit) a threshold (Image > Adjust > Threshold) and a mask of LAMP1 particles were created (Process > Binary > Convert to mask), and overlapping particles were distinguished (Process > Binary > Watershed). Finally, LAMP 1 particles were counted for each of the 4 rings (Analyze > Analyze particles), with the number counted for the ROI defined by the outermost ring representing 100%. The data was expressed as the % of total LAMP1^+^ vesicles present within each ring, thus representing the dispersion of endolysosomes in the perinuclear region (nucleus proximal rings 1 and 2) towards the cell periphery (nucleus distal rings 3 and 4).

### Autophagic flux assay

200,000 fibroblasts per well of a 6-well were plated, and after 48 hrs conditioning treated for 5h with 20nM Bafilomycin A1 (Cayman, Cat. no. 11038) or DMSO as vehicle control. Fibroblasts were harvested in RIPA and analyzed by western blotting.

### Transmission electron microscopy

Fibroblasts were pelleted via 300 x g, 3 min centrifugation, and the pellet was fixed with 2.5% glutaraldehyde, and 0.1M cacodylate buffer for 1 h at room temperature. The fixed pellet was dislodged and embedded into 3% UltraPure LMP Agarose (Invitrogen, Cat. no. 16520-100) diluted in water for 1 h at room temperature. The agarose-embedded pellet was further fixed overnight at 4°C. Transmission electron microscopy (TEM) processing was performed at the Center for Advanced Microscopy part of the Robert H. Lurie Comprehensive Cancer Center, Northwestern University, and, for imaging, the CAM/FEI Spirit G2 TEM microscope was used. Autophagic structures and endolysosomes were classified according to previously established morphological criteria^52^

### Assessment of lysosomal GCase activity via flow cytometry

The protocol used was adapted from a published protocol assessing lysosomal GCase in human monocytes.^53^ Fibroblasts were plated at the density of 100,000 fibroblasts per well of a 12-well plate for 48 h. Following trypsin dissociation, cells were pelleted by 500xg, 3 min, RT centrifugation and resuspended in 100 µL of fibroblast medium containing 250 µM PFB-FDGlu substrate (Cat. no. P11947) for 30 min in a 37°C incubator. The reaction was stopped by the addition of 1 mL ice-cold FACS buffer ((DPBS Ca^2+^/Mg^2+^-free (Gibco, 14190-144), with 1 mM EDTA (Invitrogen, 15575-020), 25 mM HEPES (Sigma, H0887), and 5% v/v HI-FBS). The cell suspension was transferred into flow cytometry tubes followed by 500xg, 3 min, RT centrifugation, and washed twice with an additional 1 mL FACS buffer. After the second wash, cells were gently resuspended in a 300 µL ice-cold FACS buffer. The green-fluorescent PFB-Glu has excitation/emission maxima ∼492/516 nm, and measured in Alexa488 via the instrument 3C.A1 LSR Fortessa 1 Analyzer.

### Flow cytometry assessment of DQ BSA

Fibroblasts were plated on 12-well plates at a density of 100,000 cells per well. After 72 hr conditioning, the medium was replaced by 1mL of fresh medium containing 25µg/mL DQ Red BSA (Invitrogen, D12051) or DQ Green BSA (Invitrogen, D12050) for 5.5hours, at 37°C. After the incubation, the medium was aspirated, followed by incubation with 500 µL trypsin for 5 min, at 37°C. The dissociation was stopped by the addition of 1mL ice-cold FACS buffer, centrifugation at 500xg, 3 min, RT, and followed by an additional wash with final resuspension to 300 µL ice-cold FACS buffer.

### Exosome/extracellular vesicle isolation by ultracentrifugation

All centrifugations as well as the 0.20 µm filtration were performed at room temperature, while the ultracentrifugation was performed at 4°C. Fibroblasts were cultured in a 15-cm dish until 80% confluency. Culture medium was replaced with 18 mL DMEM (Gibco, 11995-065) supplemented with 10% exosome-depleted FBS (Gibco, A27208-03) and conditioned for 24h. The conditioned medium was pre-cleared by sequential centrifugation at 300 × g, 5 min, then at 3000 × g, 10 min, and subsequently filtered through a 28-μm syringe 0.2-μm filter (Corning, 431219). Finally, 4 mL of the pre-cleared medium (corresponding to approximately 8×10^6^ cells) was ultracentrifuged (open-top thin-wall ultra-clear tube, 11 × 60 mm; Beckman Coulter, 344062) for 90 min, using the SW60 Ti rotor (Beckman Coulter). The final pellet (EV fraction) was resuspended in 30 µL of sample buffer (32.4 mM Tris HCl pH 6.8, 13.15% glycerol, 1.05% SDS).

### Generation of induced pluripotent stem cells and neuronal differentiation

Two patient-derived fibroblast lines from subjects F-I:1 and F-1:2 were reprogrammed by the Stem Cell Core Facility at Northwestern University to generate induced pluripotent stem cells (iPSCs). iPSC lines were shown to express the pluripotency markers Nanog, Oct4, SSEA-4, and Tra-1–81 through immunofluorescence analysis, and g-band karyotype analysis was performed by Cell Line Genetics (https://www.clgenetics.com/). Additionally, an iPSC isogenic BORCS5 knock-out (BORCS5-KO) line was generated using the CRISPR/Cas9 system. Optimal CRISPR guides were chosen using the CRISPR design tool^54^ to target *BORCS5* exon 2 and introduce a bi-allelic frameshift variant which would result in a complete loss of BORCS5. Guide RNAs were cloned into a plasmid expressing the Cas9 D10A nickase pSpCas9n(BB)-2A-GFP (PX461; Addgene # 48140) and Sanger sequenced to ensure proper cloning. iPSC colonies grown on a 10 cm dish were dissociated using Accutase and 5 million cells were transduced with 3 µg CRISPR guides using the Neon transfection system (Thermo Fisher). Transduced cells were then plated using mTeSR with 10 µM ROCK inhibitor. After 48 h, GFP-positive cells were sorted and plated at clonal density (10,000 cells/plate) on Matrigel-coated 10 cm dishes. Individual colonies were manually passaged and plated in 48 well plates. Clones were grown to confluence and passaged using Accutase. About 15% of cells were replated and the remaining were used for Sanger sequencing reactions. Crude genomic DNA was obtained using Viagen extraction reagents. Corrected clones were expanded, resequenced, and submitted for g-band karyotype analysis (Cell Line Genetics). The presence of pluripotency markers and a normal karyotype was confirmed in all four lines. The introduction of bi-allelic frameshift variants in BORCS5 exon 2 was demonstrated by Sanger sequencing and the complete KO of BORCS5 was confirmed by Western Blot analysis.

BORCS5 WT, BORCS5 KO, and patient-derived *BORCS5*-mutant iPSCs were differentiated into forebrain glutamatergic neurons using the previously described Ngn2-overexpression protocol.^24^ These neurons have been previously described to contain >98% MAP2-and vGlut-positive cells, express cortical neuron markers, and form functional synapses at day 21.^24^

### Live-cell confocal microscopy of iPSC-derived neuron

Confocal live-cell imaging was performed using a Nikon W1 Spinning Disk microscope with a 100×-oil objective (TIRF 100x 1.49 NA; Nikon Plan Apo). Cortical neurons were cultured and transduced or stained as described above and imaged in four-chamber glass-bottom dishes (D35C4-20-1.5-N; Cellvis) in a temperature-controlled (37°C) and a humidified chamber with 5% CO_2_. Images were acquired in single-camera mode with 500-ms exposure time. Cells were imaged at 1 frame every 2 s for 3 min total. Live cells were imaged in a temperature-controlled chamber (37°C) at 5% CO_2_ at one frame every 2–3 s. Dual-color videos were acquired as consecutive green-red images and tricolor videos were acquired as consecutive green-red-blue images. For live-cell imaging microscopy, cortical neurons were incubated with LysoTracker Red DND-99 (L7528; Thermo Fisher Scientific) (50 nM), Magic Red cathepsin B (ICT937; BioRad) (1:2,000), or Calcein AM-488 (20 nM) for 30 min in fresh culture media. Cells were imaged after three quick washes in fresh culturing media. Imaged cells were randomly selected based on Calcein-488 staining to achieve blinding of the investigator.

### Image processing and analysis

Confocal live-cell images were processed and analyzed using NIS-Elements 5.3 software (Nikon). To correct for photobleaching, time-lapsed images were corrected using intensity equalization over time and further processed using Denoise AI and the 2D deconvolution module (automatic mode). The General Analyzer tool was used for the analysis of Magic Red cathepsin B and LysoTracker Red DND-99 staining. Briefly, background correction was performed using rolling ball correction, and Magic Red and LysoTracker signal was thresholded and filtered for all objects between 0.1 and 2 µm. The number of positive puncta (MagicRed or LysoTracker Red) was calculated by the number of puncta divided by the cell area, and the mean fluorescence intensity was measured per thresholded object.

### Zebrafish husbandry

Adult wild-type (WT) zebrafish (Danio rerio; AB strain) were maintained at 28°C under a 12 h light/12 h dark cycle, according to the Westerfield zebrafish book. All zebrafish in this study used for cross-breeding were housed in groups and fed twice daily with a standardized diet of Skretting® Gemma Micro starting as of 5 days post-fertilization (dpf). Embryos were maintained at 28.5°C, collected, and staged as previously described.^55^ All experiments were conducted in accordance with the guidelines of the Canadian Council for Animal Care and approved by the Institutional Animal Care and Use Committee of INRS-LNBE.

### Generation of borcs5 F0 KO zebrafish model and rescue experiments

Zebrafish *borcs5* F0 knockout (KO) embryos were generated using the CRISPR-Cas9 system and three selected guide RNA (gRNA) target sequences, as described previously^17^. RNA synthesis and mix injection was performed following the previously described protocol. 1 nl of the mix was injected into one-cell stage embryos. Genotyping of *borcs5* mutants was performed using high-resolution melting (HRM) analysis and Sanger sequencing. Primer sequences used for genotyping and sequencing are available upon request.

For rescue and overexpression experiments in zebrafish *borcs5* F0 knockout (KO) models, complementary DNA (cDNA) constructs encoding wild-type (WT) human *BORCS5*, as well as the homozygous missense variants c.284G>A; p.(R95Q) and 296A>C; (p. H99P). Wild-type (WT) BORCS5, R95Q BORCS5 and H99P BORCS5 were subcloned into the pcs2+ vector, and mRNA synthesis was carried out using the mMESSAGE mMACHINE SP6 transcription kit (Ambion). Capped and polyadenylated mRNA of WT *BORCS5* and the mutant variants were synthesized in vitro using the mMESSAGE mMACHINE kit (Ambion). One-cell stage embryos were injected with 1 nl of wild-type *BORCS5* mRNA or mutant mRNAs (40 ng/μl) co-injected with the Cas9/gRNA mix for functional rescue in *borcs5* KO embryos. Morphological analysis of zebrafish embryos was performed under a Leica S6E stereoscope.

### Zebrafish behavioral assays

Larvae (5 dpf) were transferred individually into a 96-well plate containing 200 μl of E3 medium. The well plate was placed in the Daniovision® recording chamber (Noldus) for 30 min before the start of the experiment. Locomotor activity for 2 hours was recorded using a Basler GenIcam camera. Analysis was performed using the Ethovision XT 12 software (Noldus) to quantify the cumulative distance swam and swim velocity.

### Zebrafish phalloidin staining

To visualize muscles using phalloidin staining, 3 dpf embryos were fixed overnight at 4 °C in 4% paraformaldehyde (PFA). Phalloidin staining was performed following the previously described protocol.^17^ Analyses were conducted on Z-stack images acquired with a Zeiss LSM 780 confocal microscope.

### Zebrafish motor axon visualization

Immunohistochemical analyses were performed on 3 dpf zebrafish embryos to visualize motor neuron axonal projections. The embryos were fixed overnight at 4 °C in Dent’s fixative (20% DMSO and 80% methanol). Phalloidin staining was carried out as previously described.^17^ Analyses were conducted on Z-stack images acquired using a Zeiss LSM 780 confocal microscope.

### H&E brain staining

For brain section staining, hematoxylin and eosin (H&E) staining was performed on 5-μm paraffin-embedded brain sections of 3 dpf larvae. Sections were post-fixed in 10% formalin (Chaptec) for 5 minutes and subsequently rinsed with tap water. Tissue sections were subjected to hematoxylin staining (StatLab) for a duration of 4 minutes, followed by differentiation in an acid-alcohol solution and thorough rinsing with tap water. To enhance nuclear contrast, sections were subsequently immersed in a saturated lithium carbonate solution for 10 seconds and rinsed again with tap water. Counterstaining was performed using Eosin Y (StatLab) for 2 minutes. Finally, sections were mounted under coverslips using Permount mounting medium (Thermo Fisher) to ensure long-term preservation and optical clarity.

### p-MAPK/ERK staining

PTZ treatments were performed in the dark for 15 or 30 minutes on larvae (4 dpf), then rapidly fixed in 4% PFA and kept at 4°C overnight. Following fixation, larvae were extensively washed with phosphate-buffered saline (PBS) containing 0.1% Tween 20 and subsequently incubated in 100% acetone for 15 minutes. The acetone was then removed by washing with PBS containing 0.3% Triton X-100, followed by PBS-DT (PBS supplemented with 1% bovine serum albumin [BSA], 1% dimethyl sulfoxide [DMSO], and 1% Triton X-100). To minimize nonspecific binding, samples were blocked for 1 hour in PBS-DT supplemented with 5% normal goat serum. Larvae were then incubated overnight at 4°C with the primary antibody against phospho-MAPK1/ERK2-MAPK3/ERK1 (1:500; Cell Signaling Technology, 4370S). After multiple washes in PBS-DT, samples were incubated overnight at 4°C with the Alexa Fluor 488-conjugated goat anti-rabbit secondary antibody (1:1000; Invitrogen, A-11008). The larvae heads were subsequently mounted ventrally on slides using Fluoromount (ThermoFisher) and imaged using a Zeiss LSM 780 confocal microscope. Image processing and analysis were performed using Fiji (ImageJ).

### Tyrosine hydroxylase staining

Visualization of dopaminergic neurons was performed on 3 dpf larvae. Briefly, animals were fixed in 4% paraformaldehyde overnight at 4°C. After fixation, the larvae were rinsed several times for 1 h with PBS-Tween (0.1%) and then incubated in PBS-Tween (1%) for 2 h. Larvae were incubated in freshly blocking solution (2% normal goat serum, 1% BSA, 1% DMSO, 1% Triton-X in PBS). Then incubated in prepared blocking solution containing primary antibody TH (aTH, 1:200, Developmental Studies Hybridoma Bank) overnight at 4°C. The primary antibody was washed several times for 1 h with PBS-Tween (0.1%) and larvae were incubated in blocking solution containing an Alexa-Fluor-488-conjugated secondary antibody (1:1000, A-21042, Invitrogen) overnight at 4°C. The following day, the larvae were washed several times with PBS-Tween (0.1%) and mounted on a glass slide in Fluoromount (ThermoFisher). Slides were blinded for Z-stack imaging with a Zeiss LSM780 confocal microscope (Carl Zeiss). The images were then processed with ZEN software (Carl Zeiss).

## Statistical Analysis

Comprehensive information for statistical analysis and post-hoc tested applied is provided separately in each figure legend.

## Data Availability

All data produced in the present study are available upon reasonable request to the authors

## Acknowledgements

We would like to thank all the families described in this manuscript whose participation made this research possible.

This work was supported by the National Institutes of Health [grant 1K08NS131581 to N.E.M; grants R37 NS096241, and R35 NS122257 to D.K., and Intramural Program of NICHD project ZIA HD001607 to J.S.B.]; by the Canadian Institutes of Health Research (CIHR, OGB-177940 to S.A.P) and by the Parkinson’s Foundation (PF-SPE-874858 to N.E.M.). A.P. is supported by a Fonds de Recherche du Québec-Santé (FRQS) Doctoral scholarship. M.S. received grants from the Deutsche Forschungsgemeinschaft (SCHW866/6-1 and 7-1).

## Author contribution

N.E.M, G.M., S.A.P., J.S.B, K.P.B, D.K. contributed to the conception and design of the study. N.E.M., G.M., R.M., R.D.P., A.P., P.S., D.C., W.J.P., F.M., D.C., S.H.E., J.B., T.M., J.V., J.D.O.E., L.M., M.G.C., I.M.W., E.J.K., M.S.Z., A.S., G.Z., Z.N.A.H., E.M., S.S., L.S.M., S.H.C.A., M.S., M.S., H.H., S.A.P., J.S.B., K.P.B., D.K. contributed to the acquisition and analysis of data.

N.E.M, G.M., R.D.P., A.P., P.S, M.S., S.A.P., J.S.B, D.K. contributed to drafting the text and preparing the figures

---

**Table 1.** Summary of genetic and clinical findings of children and adult patients with bi-allelic *BORCS5* variants.

| Patients: Families ID | F-I:1 | F-I:2 | F-II:1 | F-III:1 | F-III:2 | F-IV:1 |
| --- | --- | --- | --- | --- | --- | --- |
| Coding sequence change | c.284G>A and c.382 383delCT | c.284G>A and c.382 383delCT | c.296A>C | c.284G>A | c.284G>A | c.203-1G>T |
| Amino Acid Change | p.R95Q and p.L128Vfs*86 | p.R95Q and p.L128Vfs*86 | p.H99P | p.R95Q | p.R95Q | p.? |
| Zygosity | Compound heterozygous | Compound heterozygous | Homozygous | Homozygous | Homozygous | Homozygous |
| Failure-to-thrive | N/A | N/A | Yes | Yes | Yes | N/A |
| Global developmental delay | Yes | Yes | Yes | Yes | Yes | Yes |
| Degree of intellectual disability | Profound | Profound | Profound | Profound | Profound | Profound |
| Regression | Yes | Yes | Yes | No | No | N/A |
| Spasticity | Severe | Severe | Severe | Severe | Severe | Severe |
| Hyperreflexia and upgoing toes | Yes | Yes | Yes | Yes | Yes | Yes |
| Seizures | Yes | Yes | Yes | Yes | Yes | Yes |
| Anarthria | Yes | Yes | Yes | Yes | Yes | Yes |
| Axial hypotonia | No | No | Yes | No | No | Yes |
| Parkinsonism | Yes, hypomimia and bradykinesia | Yes, hypomimia and bradykinesia | Yes, severe bradykinesia | No | No | No |
| Dystonia | Generalized dystonia with dystonic spasms | Generalized dystonia with dystonic spasms | Generalized dystonia with dystonic spasms | No | No | No |
| Limb contractures | Yes | Yes | No | Yes | Yes | Yes |
| Muscle atrophy | Yes | Yes | No | Yes | Yes | Yes |
| Peripheral neuropathy | Normal EMG/NCS | N/A | Sensorimotor demyelinating neuropathy | N/A | N/A | No |
| Swallowing difficulties | Yes, requiring PEG | Yes | Yes, requiring PEG | Yes | Yes | Yes |
| Ophthalmologic findings | Optic nerve atrophy, strabismus, poor tracking. | Optic nerve atrophy, strabismus, poor tracking. | Optic nerve atrophy. No tracking. | Optic atrophy | N/A | No tracking. Visual Evoked Potentials with absent P100 bilaterally |
| Microcephaly | Yes | Yes | Yes | Yes | Yes | Yes |
| Dysmorphic features | Yes | Yes | Yes | Yes | Yes | Yes |
| Brain MRI | Moderate-to-severe brain atrophy, marked WM volume loss with increased signal intensity on T2/FLAIR images, small T2-hypointense thalami, thinning of the CC and brainstem, mild cerebellar atrophy, small ON | Severe generalized cerebral atrophy for age, marked reduction of WM bulk with increased signal intensity on T2/FLAIR images | Severe and progressive generalized cerebral atrophy, marked WM volume loss with increased signal intensity on T2/FLAIR images, small T2-hypointense thalami, marked thinning of the CC and brainstem, mild cerebellar atrophy, small ON. Normal MR spectroscopy | Generalized cerebral atrophy, marked WM volume loss with increased signal intensity on T2/FLAIR images, small T2-hypointense thalami, thinning of the CC and brainstem, small ON. Normal MR spectroscopy. | Generalized cerebral atrophy, marked reduction of WM bulk with increased signal intensity on T2/FLAIR images, small T2-hypointense thalami. MR spectroscopy with reduced cerebral metabolites. | CC agenesis and polymicrogyria |
| Current effective treatment | L-DOPA, Baclofen, Gabapentin, Diazepam, Trihexyphenidyl | L-DOPA, Baclofen, Gabapentin, Diazepam, Trihexyphenidyl | L-DOPA, Trihexyphenidyl, Baclofen, Diazepam, Levetiracetam | Topiramate and Levetiracetam | Topiramate and Levetiracetam | N/A |

Legend: CC=corpus callosum, EMG=electromyography, N/A=Not available, NCS=nerve conduction study, ON=Optic Nerve, PEG=percutaneous endoscopic gastrostomy, WM=white matter.

## References

1. Ysselstein, D., Shulman, J.M. & Krainc, D. Emerging links between pediatric lysosomal storage diseases and adult parkinsonism. Mov Disord 34, 614–624 (2019).

2. Robak, L.A. et al. Excessive burden of lysosomal storage disorder gene variants in Parkinson’s disease. Brain 140, 3191–3203 (2017).

3. Steel, D. et al. Loss-of-Function Variants in HOPS Complex Genes VPS16 and VPS41 Cause Early Onset Dystonia Associated with Lysosomal Abnormalities. Ann Neurol 88, 867–877 (2020).

4. Calakos, N. & Zech, M. Emerging Molecular-Genetic Families in Dystonia: Endosome-Autophagosome-Lysosome and Integrated Stress Response Pathways. Mov Disord 40, 7–21 (2025).

5. Ballabio, A. & Bonifacino, J.S. Lysosomes as dynamic regulators of cell and organismal homeostasis. Nat Rev Mol Cell Biol 21, 101–118 (2020).

6. Parenti, G., Medina, D.L. & Ballabio, A. The rapidly evolving view of lysosomal storage diseases. EMBO Mol Med 13, e12836 (2021).

7. Cuddy, L.K. et al. Stress-Induced Cellular Clearance Is Mediated by the SNARE Protein ykt6 and Disrupted by α-Synuclein. Neuron 104, 869–884.e11 (2019).

8. Pu, J. et al. BORC, a multisubunit complex that regulates lysosome positioning. Dev Cell 33, 176–88 (2015).

9. Guardia, C.M., Farías, G.G., Jia, R., Pu, J. & Bonifacino, J.S. BORC Functions Upstream of Kinesins 1 and 3 to Coordinate Regional Movement of Lysosomes along Different Microtubule Tracks. Cell Rep 17, 1950–1961 (2016).

10. Rosa-Ferreira, C. & Munro, S. Arl8 and SKIP act together to link lysosomes to kinesin-1. Dev Cell 21, 1171–8 (2011).

11. De Pace, R. et al. Synaptic Vesicle Precursors and Lysosomes Are Transported by Different Mechanisms in the Axon of Mammalian Neurons. Cell Rep 31, 107775 (2020).

12. Farías, G.G., Guardia, C.M., De Pace, R., Britt, D.J. & Bonifacino, J.S. BORC/kinesin-1 ensemble drives polarized transport of lysosomes into the axon. Proc Natl Acad Sci U S A 114, E2955–E2964 (2017).

13. Snouwaert, J.N. et al. A Mutation in the Borcs7 Subunit of the Lysosome Regulatory BORC Complex Results in Motor Deficits and Dystrophic Axonopathy in Mice. Cell Rep 24, 1254–1265 (2018).

14. De Pace, R. et al. Messenger RNA transport on lysosomal vesicles maintains axonal mitochondrial homeostasis and prevents axonal degeneration. Nat Neurosci 27, 1087–1102 (2024).

15. Jia, R., Guardia, C.M., Pu, J., Chen, Y. & Bonifacino, J.S. BORC coordinates encounter and fusion of lysosomes with autophagosomes. Autophagy 13, 1648–1663 (2017).

16. Shelke, G.V., Williamson, C.D., Jarnik, M. & Bonifacino, J.S. Inhibition of endolysosome fusion increases exosome secretion. J Cell Biol 222(2023).

17. De Pace, R. et al. Biallelic BORCS8 variants cause an infantile-onset neurodegenerative disorder with altered lysosome dynamics. Brain 147, 1751–1767 (2024).

18. Charng, W.L. et al. Exome sequencing in mostly consanguineous Arab families with neurologic disease provides a high potential molecular diagnosis rate. BMC Med Genomics 9, 42 (2016).

19. Abdel-Hamid, M.S. et al. Biallelic variants in GTF3C3 encoding a subunit of the TFIIIC2 complex are associated with neurodevelopmental phenotypes in humans and zebrafish. Brain Commun 7, fcaf055 (2025).

20. Hariri, M. et al. Biogenesis of multilamellar bodies via autophagy. Mol Biol Cell 11, 255–68 (2000).

21. Minakaki, G. et al. Autophagy inhibition promotes SNCA/alpha-synuclein release and transfer via extracellular vesicles with a hybrid autophagosome-exosome-like phenotype. Autophagy 14, 98–119 (2018).

22. Miranda, A.M. et al. Neuronal lysosomal dysfunction releases exosomes harboring APP C-terminal fragments and unique lipid signatures. Nat Commun 9, 291 (2018).

23. Kugeratski, F.G. et al. Quantitative proteomics identifies the core proteome of exosomes with syntenin-1 as the highest abundant protein and a putative universal biomarker. Nat Cell Biol 23, 631–641 (2021).

24. Zhang, Y. et al. Rapid single-step induction of functional neurons from human pluripotent stem cells. Neuron 78, 785–98 (2013).

25. Duquesne, S., Nassogne, M.C., Clapuyt, P., Stouffs, K. & Sznajer, Y. Phenotype description in KIF5C gene hot-spot mutations responsible for malformations of cortical development (MCD). Eur J Med Genet 63, 103991 (2020).

26. Poirier, K. et al. Mutations in TUBG1, DYNC1H1, KIF5C and KIF2A cause malformations of cortical development and microcephaly. Nat Genet 45, 639-47 (2013).

27. de Araujo, M.E.G., Liebscher, G., Hess, M.W. & Huber, L.A. Lysosomal size matters. Traffic 21, 60–75 (2020).

28. Gabandé-Rodríguez, E., Boya, P., Labrador, V., Dotti, C.G. & Ledesma, M.D. High sphingomyelin levels induce lysosomal damage and autophagy dysfunction in Niemann Pick disease type A. Cell Death Differ 21, 864–75 (2014).

29. Saito, R. et al. A neuropathological cell model derived from Niemann-Pick disease type C patient-specific iPSCs shows disruption of the p62/SQSTM1-KEAP1-NRF2 Axis and impaired formation of neuronal networks. Mol Genet Metab Rep 28, 100784 (2021).

30. García-Sanz, P. et al. N370S-GBA1 mutation causes lysosomal cholesterol accumulation in Parkinson’s disease. Mov Disord 32, 1409–1422 (2017).

31. van der Knaap, M.S. & Bugiani, M. Leukodystrophies: a proposed classification system based on pathological changes and pathogenetic mechanisms. Acta Neuropathol 134, 351–382 (2017).

32. Kim, M.J., Jeong, H. & Krainc, D. Lysosomal ceramides regulate cathepsin B-mediated processing of saposin C and glucocerebrosidase activity. Hum Mol Genet 31, 2424–2437 (2022).

33. Arrant, A.E. et al. Impaired β-glucocerebrosidase activity and processing in frontotemporal dementia due to progranulin mutations. Acta Neuropathol Commun 7, 218 (2019).

34. Valdez, C., Ysselstein, D., Young, T.J., Zheng, J. & Krainc, D. Progranulin mutations result in impaired processing of prosaposin and reduced glucocerebrosidase activity. Hum Mol Genet 29, 716–726 (2020).

35. Nalls, M.A. et al. Identification of novel risk loci, causal insights, and heritable risk for Parkinson’s disease: a meta-analysis of genome-wide association studies. Lancet Neurol 18, 1091–1102 (2019).

36. Mazzulli, J.R. et al. Gaucher disease glucocerebrosidase and α-synuclein form a bidirectional pathogenic loop in synucleinopathies. Cell 146, 37–52 (2011).

37. Sidransky, E. & Lopez, G. The link between the GBA gene and parkinsonism. Lancet Neurol 11, 986–98 (2012).

38. Blauwendraat, C. et al. Genetic modifiers of risk and age at onset in GBA associated Parkinson’s disease and Lewy body dementia. Brain 143, 234–248 (2020).

39. Magrinelli, F. et al. Dissecting the Phenotype and Genotype of PLA2G6-Related Parkinsonism. Mov Disord 37, 148–161 (2022).

40. Morgan, N.V. et al. PLA2G6, encoding a phospholipase A2, is mutated in neurodegenerative disorders with high brain iron. Nat Genet 38, 752–4 (2006).

41. Paisán-Ruiz, C. et al. Widespread Lewy body and tau accumulation in childhood and adult onset dystonia-parkinsonism cases with PLA2G6 mutations. Neurobiol Aging 33, 814–23 (2012).

42. Riku, Y. et al. Extensive aggregation of α-synuclein and tau in juvenile-onset neuroaxonal dystrophy: an autopsied individual with a novel mutation in the PLA2G6 gene-splicing site. Acta Neuropathol Commun 1, 12 (2013).

43. Monfrini, E. et al. A Novel Homozygous VPS11 Variant May Cause Generalized Dystonia. Ann Neurol (2021).

44. Monfrini, E. et al. HOPS-associated neurological disorders (HOPSANDs): linking endolysosomal dysfunction to the pathogenesis of dystonia. Brain 144, 2610–2615 (2021).

45. Sanderson, L.E. et al. Bi-allelic variants in HOPS complex subunit VPS41 cause cerebellar ataxia and abnormal membrane trafficking. Brain 144, 769–780 (2021).

46. van der Welle, R.E.N. et al. Neurodegenerative VPS41 variants inhibit HOPS function and mTORC1-dependent TFEB/TFE3 regulation. EMBO Mol Med 13, e13258 (2021).

47. Yıldız, Y. et al. Homozygous missense VPS16 variant is associated with a novel disease, resembling mucopolysaccharidosis-plus syndrome in two siblings. Clin Genet (2021).

48. Le Fevre, A. et al. Compound heterozygous Pkd1l1 variants in a family with two fetuses affected by heterotaxy and complex Chd. Eur J Med Genet 63, 103657 (2020).

49. Retterer, K. et al. Clinical application of whole-exome sequencing across clinical indications. Genet Med 18, 696–704 (2016).

50. Mencacci, N.E. et al. De Novo Mutations in PDE10A Cause Childhood-Onset Chorea with Bilateral Striatal Lesions. Am J Hum Genet 98, 763–71 (2016).

51. do Couto, N.F., Queiroz-Oliveira, T., Horta, M.F., Castro-Gomes, T. & Andrade, L.O. Measuring Intracellular Vesicle Density and Dispersion Using Fluorescence Microscopy and ImageJ/FIJI. Bio Protoc 10, e3703 (2020).

52. Eskelinen, E.L. Maturation of autophagic vacuoles in Mammalian cells. Autophagy 1, 1–10 (2005).

53. Hughes, L.P., Halliday, G.M. & Dzamko, N. Flow Cytometry Measurement of Glucocerebrosidase Activity in Human Monocytes. Bio-protocol 10, e3572 (2020).

54. Haeussler, M. et al. Evaluation of off-target and on-target scoring algorithms and integration into the guide RNA selection tool CRISPOR. Genome Biol 17, 148 (2016).

55. Kimmel, C.B., Ballard, W.W., Kimmel, S.R., Ullmann, B. & Schilling, T.F. Stages of embryonic development of the zebrafish. Dev Dyn 203, 253–310 (1995).

