## Supplemental material for "Pathogenic variants in *BORCS5* Cause a Spectrum of Neurodevelopmental and Neurodegenerative Disorders with Lysosomal Dysfunction"

Supplementary table 1. Details of identified *BORCS5* variants

| **Family** | **Chromosome position (GRCh38)** | **cDNA (ENST00000314565.9; NM_058169)** | **Amino acid change** | **GnomAD MAF** | **Igenomix Database MAF** | **UCL Queen Square Genomics Database MAF** | **CADD** | **Polyphen-2** | **SIFT** | **Mutation Taster** | **SpliceAI** |
|---|---|---|---|---|---|---|---|---|---|---|---|
| F-IV | 12-12435627-G-T | c.203-1G>T | p.? | 0 | 0 | 0 | 35 | N/A | N/A | N/A | 1.00 (Acceptor Loss) |
| F-I, F-III | 12-12435709-G-A | c.284G>A | p.R95Q | 0.000003717 (6 het carriers) | 0 | 0 | 29.5 | probably_damaging | deleterious | disease_causing | 0 |
| F-II | 12-12435721-A-C | c.296A>C | p.H99P | 0 | 0 | 0 | 26.4 | probably_damaging | deleterious | disease_causing | 0 |
| F-I | 12-12465564-ACT-A | c.380_381delCT | p.L128Vfs*86 | 0.00001115 (18 het carriers) | 0.00000792921 (1 HET carrier) | 0 | N/A | N/A | N/A | N/A | 0 |
| F-V, F-VII | 12-12435740-GG-G | c.316delG | p.A106Pfs*20 | 0 | 0 | 0 | N/A | N/A | N/A | N/A | 0 |
| F-VI | 12-12465602-C-G | c.417C>G | p.Y139* | 0.000004337 (7 het carriers) | 0.0000158587 (2 HET carriers) | 0 | 33 | N/A | N/A | N/A | 0 |

MAF=minor allele frequency; HET=heterozygous; N/A=not applicable

**Supplementary figures**

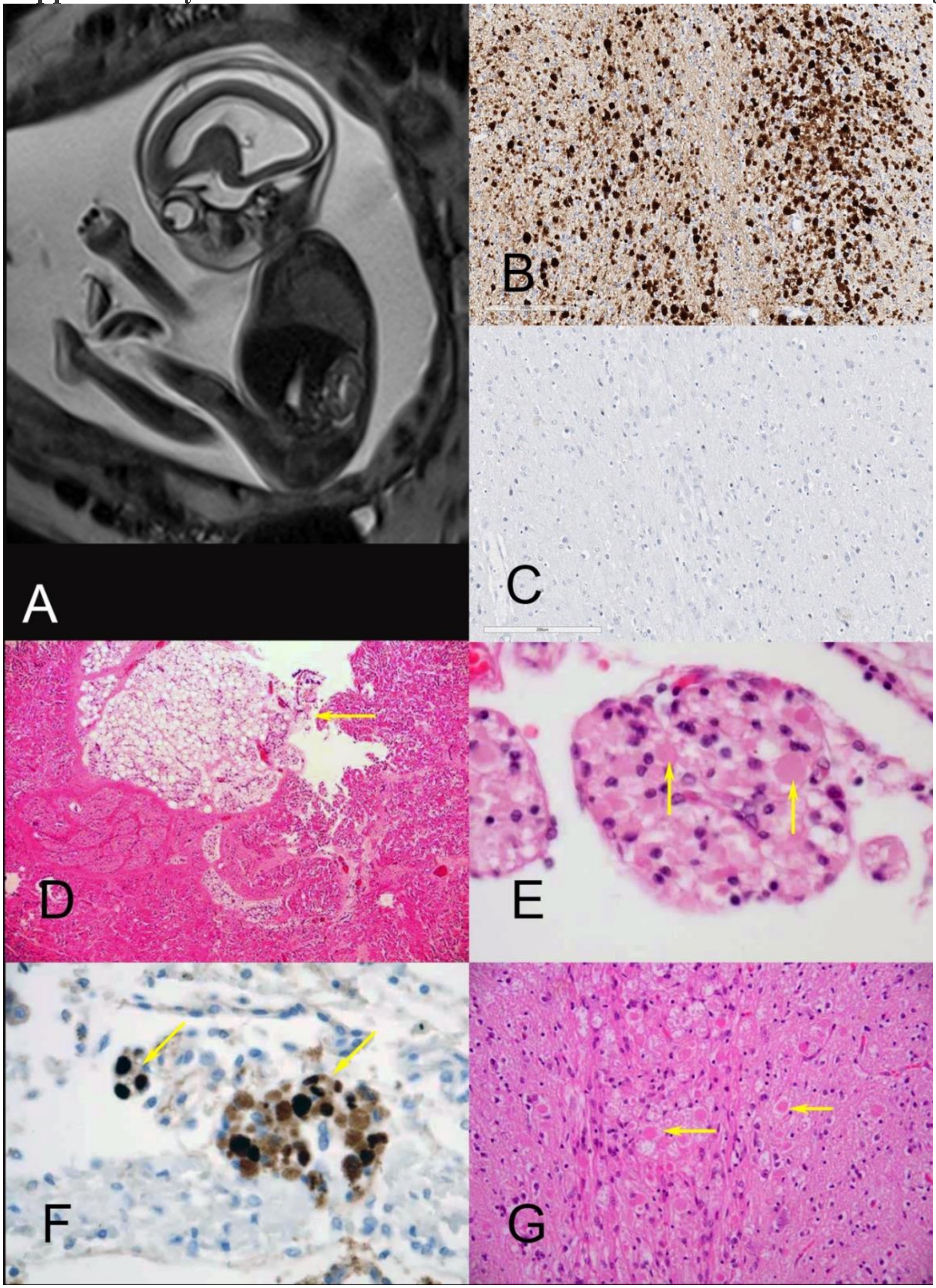


**Supplementary Figure 1.** A. MRI Sagittal T2-weighted image of the fetus F-VII:1 showing markedly reduced fetal motion with hyper extended extremities and generalized decreased muscle bulk consistent with arthrogryposis multiplex congenita (AMC). B. Internal capsule of the fetus F-VII:1 with innumerable axonal swellings staining for beta amyloid precursor protein. C. Negative staining for aggregated phosphorylated alpha synuclein. D. Skeletal muscle of case F-V:2. demonstrating small myofascicles with excess fibromyxoid perimysium and fat (arrow). E-F. Cranial nerve root of case F-V:2 containing many large eosinophilic axonal spheroids (arrow) (E), which stained positive for β-amyloid precursor protein (arrows) (F). G. Brainstem of case F-V:2 showing widespread eosinophilic axonal spheroids (arrows).

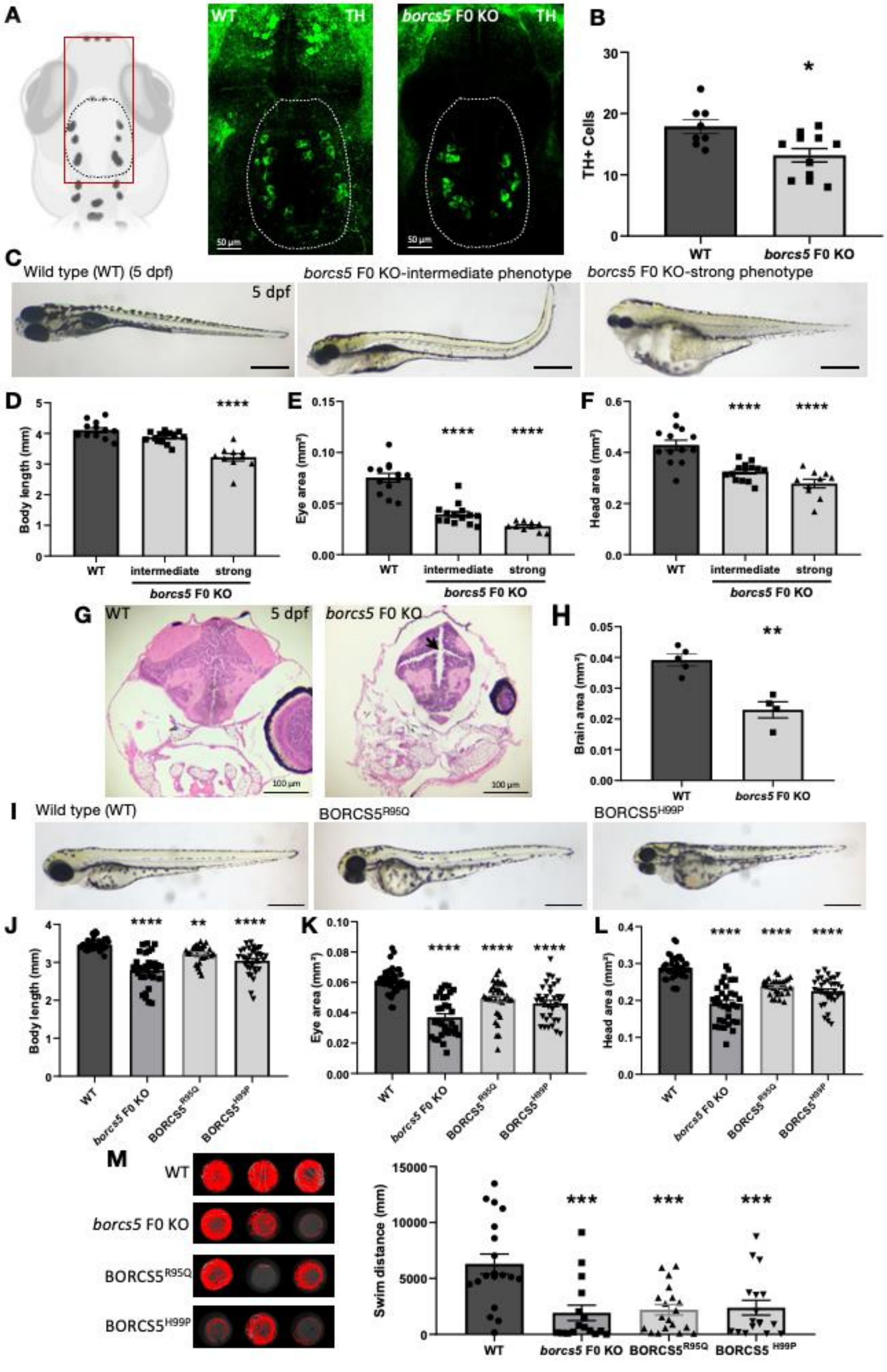

A
WT
TH
borcs5 F0 KO
TH
50 μm
50 μm
B
30
20
10
0
TH+ Cells
*
WT
borcs5 F0 KO
C
Wild type (WT) (5 dpf)
5 dpf
borcs5 F0 KO-intermediate phenotype
borcs5 F0 KO-strong phenotype
D
Body length (mm)
****
WT
intermediate
strong
borcs5 F0 KO
E
Eye area (mm²)
**** ****
WT
intermediate
strong
borcs5 F0 KO
F
Head area (mm²)
**** ****
WT
intermediate
strong
borcs5 F0 KO
G
WT
5 dpf
borcs5 F0 KO
100 μm
100 μm
H
Brain area (mm²)
**
WT
borcs5 F0 KO
I
Wild type (WT)
BORCS5R95Q
BORCS5H99P
J
Body length (mm)
**** ** ****
WT
borcs5 F0 KO
BORCS5R95Q
BORCS5H99P
K
Eye area (mm²)
**** **** ****
WT
borcs5 F0 KO
BORCS5R95Q
BORCS5H99P
L
Head area (mm²)
**** **** ****
WT
borcs5 F0 KO
BORCS5R95Q
BORCS5H99P
M
WT
borcs5 F0 KO
BORCS5R95Q
BORCS5H99P
Swim distance (mm)
*** *** ***
WT
borcs5 F0 KO
BORCS5R95Q
BORCS5 H99P

**Supplementary figure 2.** A. Tyrosine hydroxylase (TH) staining dopaminergic neurons at 3 dpf of WT and borcs5-ko larvae. Scale bars: 50 µm. B. Quantification of tyrosine hydroxylase (TH) positive neurons in diencephalic (paraventricular organ) region for WT, *borcs5* F0 KO (N=2, n=8-11). C. Morphology of zebrafish WT, and borcs5-ko larvae at 5 dpf. Scale bars: 500 µm. (D-F) Body length, eye size, and head size of WT, borcs5-ko larvae at 5 dpf (N=2, n=10-14). G. Midbrain sections stain with hematoxylin & eosin of larvae of 5 dpf. Scale bar: 100 µm. The arrow indicates ventriculomegaly in borcs5-ko larvae. H. Brain area comparison of borcs5-ko larvae (N=4) relative to WT (N=5). I. Morphology of zebrafish WT and variants models, $BORCS5^{R95Q}$ and $BOCRS5^{H99P}$ larvae at 3 dpf. Scale bars: 500 µm. (J-L) Body length, eye size, and head size of WT (N=3, n=40), borcs5-ko (N=3, n=32), $BORCS5^{R95Q}$ (N=2, n=32-35) and $BORCS5^{H99P}$ (N=2, n=34) larvae at 3 dpf. (M) $BORCS5^{R95Q}$ (n=19) and $BORCS5^{H99P}$ (n=19) larvae show motor behavior comparable to borcs5-ko (n=19) and display impaired swim distance and velocity compared to WT (n=19). All data are represented as the mean ± SEM. Statistical significance was calculated by one-way ANOVA followed by Tukey's multiple comparisons tests, or Student's T-test. *P < 0.05; **P < 0.01; ****P < 0.0001.

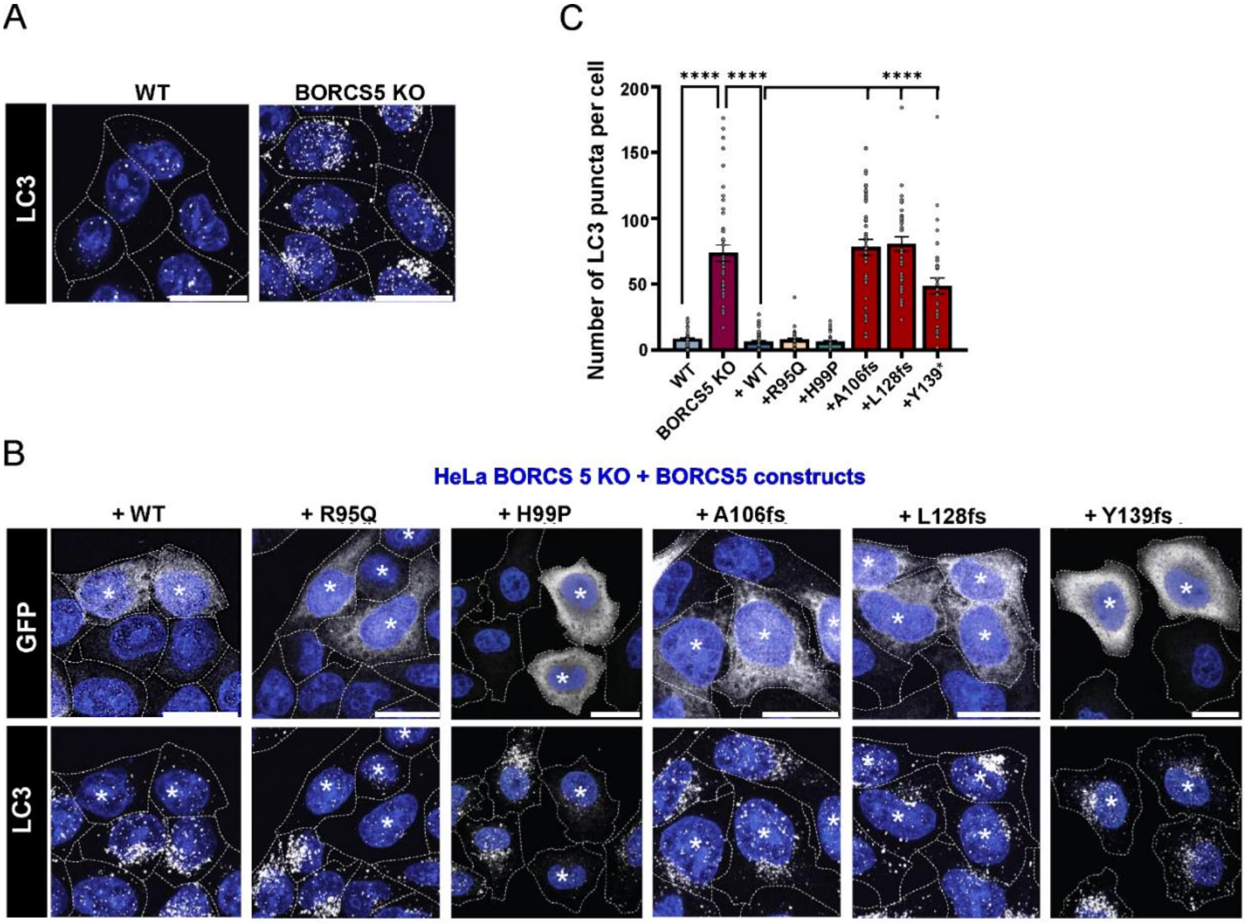


**Supplementary figure 3:** A-B. ICC shows endogenous LC3 (white puncta) distribution in untransfected WT and BORCS5 KO HeLa cells as control. BORCS5 KO HeLa cells were transiently co-transfected with the indicated BORCS5 constructs and GFP. ICC shows endogenous LC3 distribution in GFP+ transfected cells (indicated by asterisk). Nuclei were labeled with DAPI (blue), and cell edges were outlined by fluorescent phalloidin (indicated by dashed lines). Scale bars: 20 µm. C. Quantification shows mean±SEM, N=3 independent experiments. Statistics: One-way ANOVA with Tukey's multiple comparisons test (mean of each column compared to the mean of every other column), $F_{LC3\ puncta}(7,320)=72.53$, $P<0.0001$. ****$p < 0.0001$.

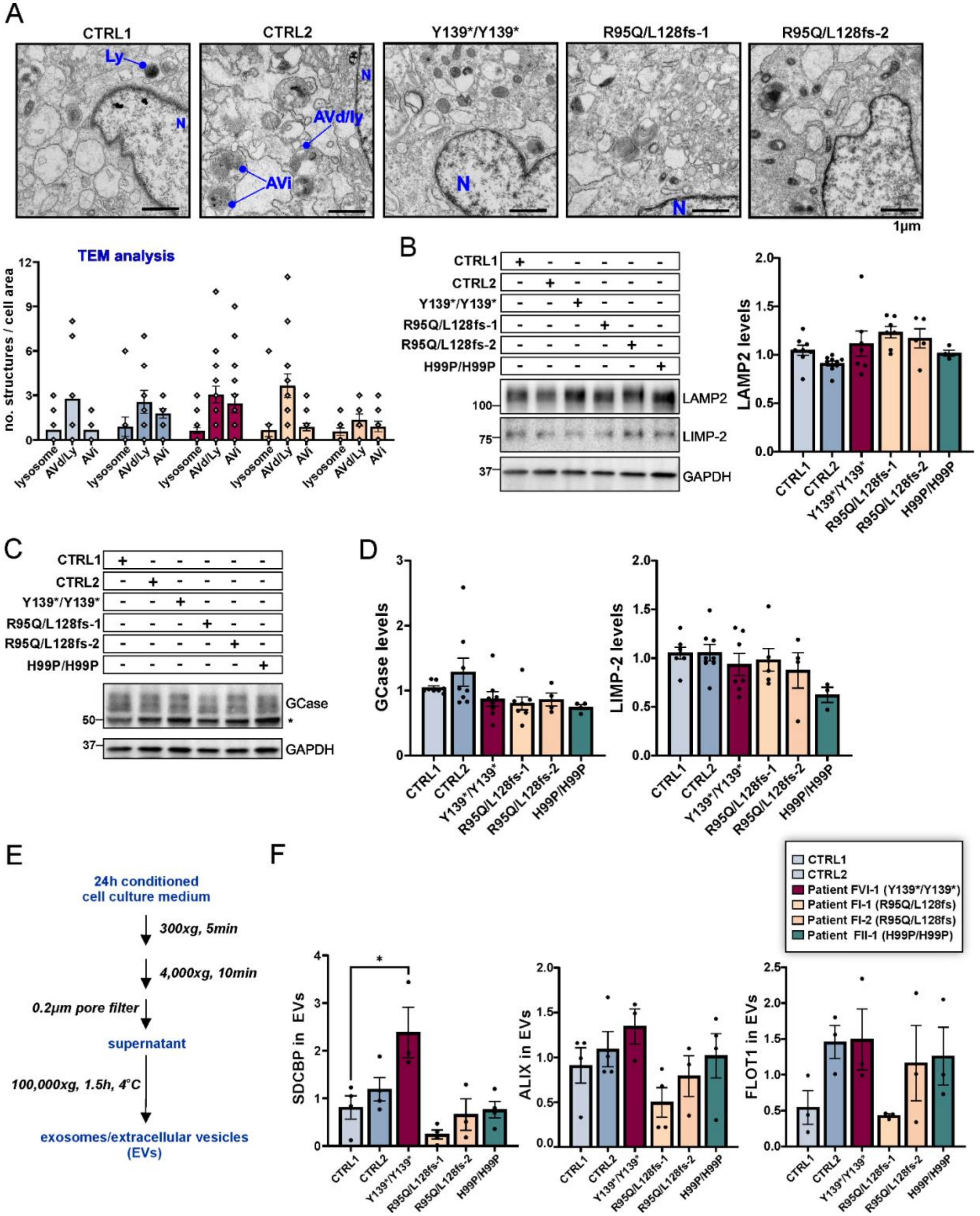


**Supplementary figure 4:** A. TEM of fibroblasts from the indicated *BORCS5* genotypes. Individual autophagic structures were classified according to previously published criteria.[52] Outlined insets are presented at higher magnification on the right, indicated by dashed lines.

Abbreviations: N: Nucleus; Avi: Early/initial autophagic vacuole; Avd: Degradative autophagic vacuole/autolysosome; Ly: Lysosome. Graph shows mean±SEM of the number of individual structures identified in individual cells, N=9 to 18 individual cells per fibroblast line. B-D. WB and quantification of relative lysosomal protein amounts in the indicated control and BORCS5 patient fibroblasts. Graphs show mean±SEM over the mean of control lines, N=3-6 independent experiments. Statistics: One way ANOVA, $F_{LAMP2}(5,31) = 2.86$, P=0.031; $F_{LIMP\text{-}2}(5,26)=1.354$, P=0.274, P=0.21; $F_{GCase}(5,30)=1.794$, P=0.144. Asterisk indicates non-specific band in the GCase blot. E. Schematic representation of fibroblast EV isolation via ultracentrifugation. F. Graphs of EV protein markers (WB of Fig. 7F) show mean±SEM, N=3-4 independent experiments. Statistics: One way ANOVA with Tukey's multiple comparisons test (mean of each column compared to the mean of every other column), $F_{SDCBP}(5,16)=6.691$, P=0.0015. *p=0.011; $F_{ALIX}(5,16)=1.843$, P=0.1611; $F_{FLOT1}(5,12)=1.743$, P=0.1993.
